# The Multiethnic Cohort: A Resource for the study of Genetic and non-Genetic Cancer Risk Across Populations

**DOI:** 10.1101/2025.06.09.25328993

**Authors:** David Bogumil, Xin Sheng, Peggy Wan, Lucy Xia, Loreall Pooler, Iona Cheng, Samantha Streicher, Brian Z. Huang, Fei Chen, Daniel Stram, Sylvia Shen, Gillian King, Charleston W. K. Chiang, Chrissie Ongaco, Marcia Adams, Ivy McMullen, Peng Zhang, Hua Ling, Michelle Mawhinney, Kimberly F. Doheny, Loïc Le Marchand, Lynne R. Wilkens, Christopher A. Haiman, David V. Conti

## Abstract

**Introduction:** The Multiethnic Cohort Study (MEC) is a U.S. prospective cohort of over 215,000 participants, designed to investigate variation in risk factors and disease across diverse racial and ethnic groups. Over 74,000 participants contributed biospecimens for genetic studies. We describe this sub-cohort and demonstrate the types of analyses it enables.

**Methods:** The MEC recruited adults aged 45–75 in California and Hawaii between 1993 and 1996. Cancer diagnoses were identified via state tumor registries. The MEC Genetics Database includes 73,139 participants with germline genotype data. We evaluated genetic similarity, its relationship with self-reported race/ethnicity, and baseline characteristics, including neighborhood socioeconomic status. Using breast, colorectal, and prostate cancer as examples, the database supports multi-ancestry genome-wide association studies (GWAS), evaluation of non-genetic factors, and time-to-event analyses.

**Results:** Participants included 10,962 African Americans, 24,234 Japanese Americans, 17,242 Latinos, 5,488 Native Hawaiians, 14,649 Whites, and 564 other. Principal component analysis revealed substantial diversity in ancestry. Multiethnic GWAS demonstrated effective control of population stratification while replicating many previously discovered variants. Polygenic risk score (PRS) effects varied by racial and ethnic group. Time-to-event analysis showed associations between cancer incidence and neighborhood socioeconomic status, population descriptors, and genetic similarity.

**Discussion:** The MEC Genetics Database enables comprehensive assessment of genetic and non-genetic cancer risk, revealing differences in absolute risk by race and ethnicity. Studying both types of risk factors in diverse and admixed populations is critical for improving risk characterization and reducing disparities. This resource supports future research in polygenic traits, gene-environment interactions, and integrated risk prediction.

## Introduction

The Multiethnic Cohort Study (MEC) is a National Cancer Institute (NCI)-funded prospective cohort study that was established in Hawaii and California (mainly Los Angeles) to identify factors that underlie disease variation across several common racial and ethnic populations in the U.S.^1^ Recruitment took place between 1993 and 1996. The MEC comprises 215,251 individuals from diverse racial and ethnic backgrounds, allowing for prospective analysis of cancer and other chronic diseases in relation to various lifestyle exposures, particularly diet.^2^ The comprehensive data collection, including biological specimens and detailed questionnaires, ensures a remarkably high level of completeness, while the high level of variation in demographic characteristics enhances the generalizability of findings. Consistent with Surveillance, Epidemiology, and End Results (SEER) reports, ^3,4^ MEC investigations have documented racial and ethnic differences in cancer risk, including higher breast cancer incidence rates in Native Hawaiians and European Americans ^5^, higher colorectal cancer rates in Japanese American and African American populations ^6^, and higher prostate cancer rates in African American males.^2^

In addition to identifying these disparities, detailed questionnaire data and participant diversity unique to the MEC have allowed researchers to investigate the sources of these health disparities by estimating exposure-disease associations across racially and ethnically diverse populations. The MEC has been used to investigate differences in the effects of smoking on risk of breast cancer, lung cancer, and pancreatic cancer,^7–9^ differences in the effects of diabetes on risk of several cancers,^10,11^ the effects of diet, ^12,13^ the effect of air pollution on cancer risk^14,15^, and the influence of structural and social determinants of health (SSDoH) such as neighborhood socioeconomic (nSES) status and the built environment on cancer risk.^16,17^ In many cases, examination of common non-genetic risk factors of these cancers does not fully explain differences in risk between populations,^5,7,10^ suggesting a potential role of germline variation in disease etiology.^5^

To investigate health disparities and predict disease risk, detailed population descriptors and comprehensive data on genetic and non-genetic risk factors are essential^18–20^. Joint modeling of these variables helps epidemiologists identify independent effects of risk factors, assess heterogeneity, and understand how variation in risk factor prevalence contributes to health disparities. Unaccounted phenotypic variation and distribution differences in non-genetic risk factors can lead to heterogeneous effects across populations and influence cross-sample polygenic risk score (PRS) performance^20,21^.

Context-specific PRS risk estimation addresses many of these challenges by modeling both genetic and non-genetic factors^22^. Due to the lack of diversity in genomics research and limited data to use in joint models, current risk prediction methods may lead to an exacerbation of health disparities^23–28^.

In the MEC, over 74,000 participants contributed biological samples that have been genotyped for disease-focused studies, including, colorectal cancer, lung cancer, pancreatic cancer, prostate cancer, type 2 diabetes, adiposity, and Alzheimer’s Disease. The MEC has also contributed to several consortium analyses, replication analyses of genome-wide association study (GWAS)-discovered variants, and polygenic risk score evaluation in admixed samples across several chronic disease phenotypes.^29–33^ Much of the research in the MEC has supported the hypothesis that genetic variation is likely to explain a portion of known racial and ethnic health disparities for several chronic diseases.^34–37^

Here, we describe the development of a single, multiethnic, database of germline variation, non-genetic risk factors, and cancer incidence data. Detailed information regarding sample characteristics, genotype quality control, imputation, and genetic similarity estimation are provided to assist with and facilitate use by researchers. We illustrate the use of this resource for discovery via GWAS for multiple cancers, risk prediction via PRS in time-to-event prospective analyses, and demonstrations of the complex relationship between genetic and non-genetic variables via integrated multivariable analyses.

## Methods

### Cohort Enrollment and Case Ascertainment

The MEC is composed of over 215,000 individuals across five self-reported racial and ethnic groups (African American, Japanese American, Latino, Native Hawaiian and White) who were 45-75 years old and resided in California (mainly Los Angeles County) or Hawaii at enrollment. Participants were identified for recruitment using the Department of Motor Vehicle records, Health Care Financing Administration files, and voter registration lists and recruited into the study from 1993 through 1996^2^. Participants were mailed a package with a letter that explained study goals and contained a 26-page baseline questionnaire (English or Spanish) that collected self-reported information on diet, demographic characteristics, smoking history, anthropometric measures, reproductive factors, chronic conditions, and physical activity.^1^ The nSES of MEC participants was based on residential address at baseline. Addresses were geocoded to land parcels or street segments, then geocoded and linked to the

U.S. Census block group.^14^ Participants were excluded from nSES sub-analyses with address records at baseline that could not be geocoded to a census block group. nSES was categorized into quartiles using state distributions.

Cancer diagnoses were identified via regular linkage to the California Cancer Registry and the Hawaii Tumor Registry, which are statewide registries that are members of the National Cancer Institute’s Surveillance, Epidemiology, and End Results (SEER) Program. Vital status and cause of death are identified through regular linkage to the National Death Index and state death certificate files. For this study, prostate cancer was defined using ICD-O codes C619, breast cancer ICD-O C500-C509, and colorectal cancer ICD-O C180-C189, C260, C199, C209. All cancer types excluded cases with ICD-O histology codes 9050-9055, 9140, or 9590-9992, and included only invasive tumors (behavior > 2).

Race and ethnicity was defined by self-report in the baseline questionnaire using terminology and labeling in practice in the early 1990s when the surveys were distributed to participants. Race and ethnicity are defined in accordance with the U.S. Census Bureau in 1990, where race refers to an individual’s self-defined classification, and ethnicity refers to an individual’s self-reported Hispanic Origin. Responses were categorized into one of five groups and a single “other” group when a selection corresponding to one of the five major groups was not provided. When participants reported multiple racial/ethnic groups, the categorization procedure prioritized group membership in the following order: “African American”, “Hawaiian”, “Latino”, “Japanese”, “White”, “Filipino”, “Chinese”, “Korean”, and “Other”. Among the 0.3% of individuals where no response was given for the participant but was provided for the mother and father, a race and ethnicity value was created from the parents’ information.

The University of Hawaii and the University of Southern California Institutional Review Boards reviewed and approved this study, including that return of a completed questionnaire was considered as proof of informed consent. The review and approval from institutional review boards are in accordance with the Department of Health and Human Services regulations in 45 CFR 46.

### The MEC Biorepository and Genetics Database

Biospecimen collection in the MEC took place in two main phases. In Phase 1, blood and buccal samples were collected from incident cancer cases and controls between 1995 and 2001 for nested case-control studies of prostate, breast, and colorectal cancer (n=6,359 cancer cases and 9,193 non-cases). In Phase 2, between 2001 and 2007, MEC participants living within the catchment areas of University of Hawaii and University of Southern California were contacted to provide a biospecimen, with 60,280 providing one. All MEC participants provided signed informed consents prior to biospecimen collection. Since 2010, 17,197 specimens have been genotyped, primarily with Illumina arrays, as part of 30 GWAS studies of cancer and other chronic diseases. In 2020-2021, the remaining MEC subjects with biosamples (n=33,831) were genotyped with the Illumina Global Diversity Array (with 1,645 of the samples from Phase 1 re-genotyped in Phase 2 as quality control). The MEC Genetics Database includes biorepository participants with germline variant data (n=73,139) which passed quality control procedures (described below). The MEC Genetics Cohort is a subsample comprised of individuals who were cancer-free at biospecimen collection (e.g. excluding those with prostate, breast, and colorectal cancer recruited during phase 1; n= 65,408; Figure 1). All cancer cases within the MEC Genetics Cohort are incident cases (relative to the biospecimen collection date, including 1,880 incident breast cancer cases, 1,340 incident colorectal cancer cases, and 2,899 incident prostate cancer cases), which were used in risk estimation.

**Figure 1:**
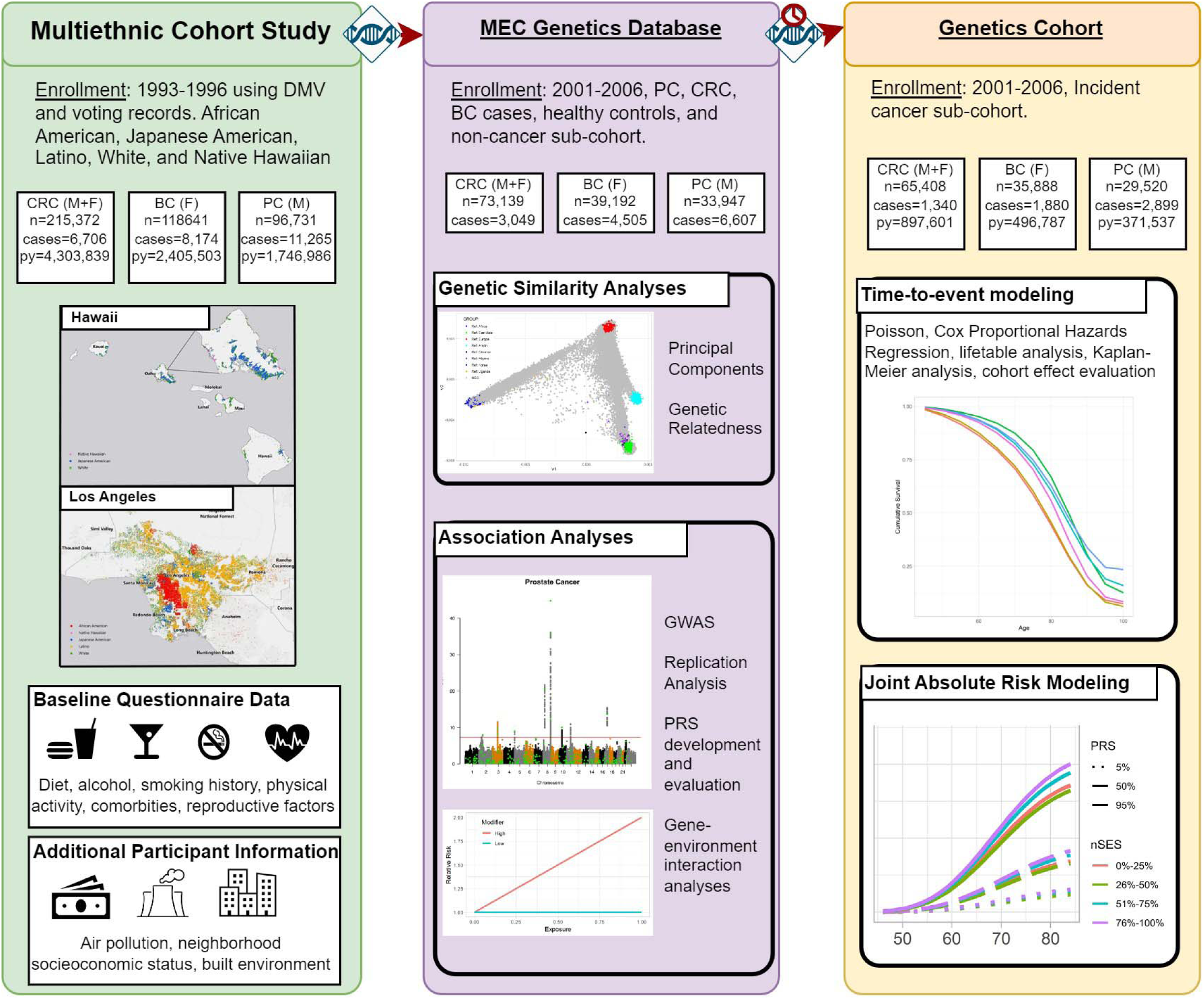
Participant Inclusion Diagram. Flow diagram showing case counts, person-years, data types available, and analytical approaches avaiable for each type of data. BC - Breast Cancer, CRC – Colorectal Cancer, PC – Prostate Cancer

### Genotyping Quality Control Process

Genotyped samples underwent an intensive quality control process of sample and variant call rate filtering of over 0.95, within race and ethnicity Hardy Weinberg Equilibrium violation testing, allele matching, and allele frequency comparisons against corresponding super populations from the 1000 Genomes Project reference data (supplementary text). Following quality control filtering, imputation was performed using the Minimac4 algorithm and the 1000 Genomes Project and TOPMed reference panels.^38–40^ Imputation was conducted within self-reported race and ethnicity and study, allowing for the calculation of imputation quality statistics within these groups. Projects that utilized the same or similar Illumina arrays, primarily for MEGA chip and OncoArray projects, were merged into a single dataset for imputation.

### Population Structure Estimation

We estimated genetic similarity using principal components and clustering within the sample and relative to external continental reference data.^41^ These variables were also used to adjust for potential confounding by population structure in genetic analyses. Common autosomal single nucleotide variants (SNVs) across the 30 genotyping platforms that were directly genotyped in all GWAS projects were identified and subsequently subjected to linkage disequilibrium (LD) pruning with an R^2^ threshold of < 0.1 using all subjects. This resulted in approximately 15.7K SNVs used to estimate principal components by PCair and PCrelate.^42,43^ We incorporated 1,890 individuals from two external data sources into our principal component analysis dataset: the 1000 Genomes Project and the PAGE Global Reference Panel (supplemental text) to anchor study participants with known reference panel individuals. ADMIXTURE was used to estimate similarity clusters among MEC participants and the 1,890 reference individuals.^41^ The unsupervised analysis used K=5 and 21,431 SNVs. These K groups are labeled as “clusters.” Compared to reference panel samples, these clusters corresponded to European-, African-, and East Asian-ancestry, with additional clusters of suspected Amerindigenous-ancestry within the MEC Latinos and Polynesian ancestry identified in the MEC Native Hawaiians. As there is ambiguity about the specific ancestry estimated, we do not label or infer any socially constructed label, such as race or ethnicity, by these groupings. We report cluster proportion across participants by self-reported race and ethnicity.

### Covariate and Population Descriptor Correlations

To illustrate the associations between self-reported race and ethnicity, genetic similarity, and baseline covariates, we used generalized linear models to estimate partial correlations between baseline covariates and principal components, adjusting for age at sample collection and sex. The model took the form *g(. ) = b*_O_ *+ b*_age_ *x*_age_ *+ b*_sex_ *x*_sex_ *+P* , where *g(.)* was the identity or logit link and *P* was either a set of 10 principal components, self-reported race and ethnicity, or both. We conducted this partial correlation analysis in the multiethnic sample and stratified within each self-reported race and ethnic group. Categorical variables were dichotomized when appropriate and modeled using logistic regression. Continuous variables were modeled using linear regression.

### Genome-wide Association Analysis

Using the MEC Genetics Database, we illustrate a multiethnic GWAS using prostate cancer (6,607 cases), breast cancer (4,505 cases), and colorectal cancer (3,049 cases) as example phenotypes, with non-cancer participants as controls (Figure 1). We conducted the GWAS using a mixed models approach implemented through SAIGE to account for cryptic relatedness between samples.^44^ GWAS models were adjusted for age at sample collection, sex, the top ten principal components, and the study project. Variants with R^2^ > 0.8 across all imputed project datasets (M=5,017,063) were analyzed; rare variants with minor allele counts of ten or fewer in the overall sample were removed.

### Genetic Risk Score Analysis

For each of the three cancer phenotypes, we estimated the association between genetic risk and incident cancer using log-odds weighted genetic risk scores in the MEC Genetics Cohort (n=65,408; 1,880 breast cancer cases; 1,340 colorectal cancer cases, 2,899 prostate cancer cases). The variants in each PRS were identified using the most current GWAS results for colorectal, prostate, and breast cancer risk.^45–47^ The weighted polygenic risk score was calculated as 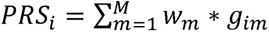, where *g_im_* is the genotype dosage for individual *i* for variant *m,* and *w*_m_ is the variant-specific log(OR) weight reported from the corresponding origional GWAS. All PRS were standardized within self-reported race and ethnicity by the PRS distribution within controls. In PRS time-to-event analysis, age at sample collection was used at entry, and age at first cancer occurrence (for each respective cancer-specific analysis) or death was used as censorship time. Using individual-level data, we conducted high-dimensional data tabulation to create Poisson distributed data with tabulated strata case counts, within strata person-years, and person-year weighted mean PRS values (supplemental text). We estimated the association between the standardized PRS (continuous variable) and incident cancer in a time-to-event analysis, using the tabulated data, with person-years as an offset term. We report hazard ratios (HR) and corresponding Wald 95% confidence intervals for all PRSs. Using the resulting Poisson models and complementary all-cause mortality time-to-event models, we estimated each cancer’s cumulative risk and 5-year risk across age and PRS quantiles (supplemental text).

We additionally explored the added contribution of nSES and genetic similarity in risk stratification within levels of PRS on the cumulative and 5-year risk scales. For nSES, these models were fit as previously described but included interaction terms between PRS and categorical nSES. This analysis allowed us to examine how much variation in risk could be attributed to nSES within levels of PRS. Lastly, we explored an approach for risk estimation that does not use self-reported race and ethnicity in risk prediction but bases risk estimation on genetic similarity across samples. For this method, we estimated cancer risk as a function of age, polygenic risk, and genetic similarity (cluster values described above). We compared predicted risk estimates against a self-reported population descriptor approach to identify differences in risk prediction across approaches.

## Results

### Sample Sizes

Participant characteristics are shown in Figure 1 and Table 1. Of the complete MEC cohort (n= 215,372), 34% (n=73,139) are included in the MEC Genetics Database, and 30% in the MEC Genetics Cohort (Figure 1, Table 1). There were no notable differences in the distributions of common baseline questionnaire covariates across the total MEC cohort, the MEC Genetics Database, and the MEC Genetics Cohort (Supplemental Figure 1). Although the differences were small, participation in the MEC Genetics Cohort was associated with slightly higher nSES, lower diabetes prevalence, and a higher prevalence of being a “never smoker.” The MEC Genetics Database includes 10,962 African American, 24,234 Japanese American, 17,242 Latino, 5,488 Native Hawaiian, 14,649 White participants, and 564 participants who did not self-report into one of the five groups (i.e. “other”). In this MEC Genetics Database sample, there were 4,505 cases of breast cancer, 3,049 colorectal cancer, and 6,607 prostate cancer. In the MEC Genetics Cohort, there were 1,880 breast cancer, 1,340 colorectal cancer, and 2,899 prostate cancer incident cases that were diagnosed after providing a biospecimen.

**Table 1:** Multiethnic Cohort Participant Characteristics.

|  | <u>Combined</u> | <u>African</u><br><u>American</u> | <u>Japanese</u><br><u>American</u> | <u>Latino</u> | <u>Native</u><br><u>Hawaiian</u> | <u>Other</u> | <u>White</u> |
| --- | --- | --- | --- | --- | --- | --- | --- |
| **MEC Complete Cohort** |  |  |  |  |  |  |  |
| n | 215372 | 34847 | 56917 | 47379 | 14502 | 12196 | 49531 |
| Age at Entry Mean (SD) | 60.4 (8.8) | 61.9 (9.0) | 61.6 (9.1) | 60.3 (7.7) | 57.1 (8.7) | 59.1 (8.5) | 59.5 (9.1) |
| Year of Entry Mean (SD) | 1994 | 1993 | 1994 | 1994 | 1994 | 1994 | 1994 |
| Female (%) | 118641 (55.1%) | 22119 (63.5%) | 29982 (52.7%) | 24599 (51.9%) | 8185 (56.4%) | 7145 (58.6%) | 26611 (53.7%) |
| **Cancer Cases** |  |  |  |  |  |  |  |
| Breast | 8174 | 1439 | 2303 | 1262 | 815 | 480 | 1875 |
| Colorectal | 6706 | 1222 | 2209 | 1301 | 394 | 275 | 1305 |
| Prostate | 11265 | 2201 | 3028 | 2604 | 578 | 510 | 2344 |
| **MEC Genetics Database** |  |  |  |  |  |  |  |
| n | 73139 | 10962 | 24234 | 17242 | 5488 | 564 | 14649 |
| Age at Entry Mean (SD) | 68.1 (8.4) | 69.5 (8.4) | 68.7 (8.7) | 68.1 (7.5) | 64.7 (8.2) | 68.2 (9.6) | 67.2 (8.5) |
| Year of Entry Mean (SD) | 2003 | 2002 | 2003 | 2003 | 2003 | 2003 | 2003 |
| Baseline Age at Entry Mean (SD) | 59.0 (8.5) | 61.2 (8.8) | 59.3 (8.7) | 59.3 (7.4) | 55.5 (7.9) | 58.9 (8.0) | 57.6 (8.6) |
| Female (%) | 39192 (53.6%) | 6305 (57.5%) | 12740 (52.6%) | 9181 (53.2%) | 3150 (57.4%) | 331 (58.7%) | 7485 (51.1%) |
| **Cancer Cases** |  |  |  |  |  |  |  |
| Breast | 4505 | 772 | 1342 | 778 | 430 | 135 | 1048 |
| Colorectal | 3049 | 558 | 1028 | 684 | 168 | 61 | 550 |
| Prostate | 6607 | 1352 | 1850 | 1591 | 330 | 117 | 1367 |
| **MEC Genetics Cohort** |  |  |  |  |  |  |  |
| n | 65408 | 9430 | 22228 | 15487 | 5069 | 296 | 12898 |
| Age at Entry Mean (SD) | 67.8 (8.4) | 69.4 (8.4) | 68.5 (8.7) | 67.8 (7.5) | 64.4 (8.2) | 64.9 (9.2) | 66.7 (8.5) |
| Year of Entry Mean (SD) | 2003 | 2002 | 2003 | 2003 | 2003 | 2000 | 2004 |
| Baseline Age at Entry Mean (SD) | 58.6 (8.4) | 60.9 (8.9) | 59.0 (8.7) | 58.9 (7.3) | 55.2 (7.8) | 58.2 (7.9) | 57.0 (8.5) |
| Female (%) | 35888 (54.9%) | 5658 (60.0%) | 11844 (53.3%) | 8603 (55.5%) | 2920 (57.6%) | 188 (63.5%) | 6675 (51.8%) |
| **Cancer Cases** |  |  |  |  |  |  |  |
| Breast | 1880 | 294 | 631 | 337 | 229 | 16 | 373 |
| Colorectal | 1340 | 243 | 489 | 304 | 77 | 4 | 223 |
| Prostate | 2899 | 538 | 1006 | 602 | 182 | 16 | 555 |

### Age of Entry

The mean age at entry into the MEC was 60.4 years. The mean age at entry into the MEC Genetics Database (i.e., age at collection) was 68.1 years, and the mean age of participants included in the MEC Genetics Cohort was 67.8 years (Table 1).

### Genetic Characteristics of Sample

There is a high degree of genetic diversity in the MEC Genetics Database. Figure 2 shows principal component plots of principal components 1-3 by race and ethnicity with external reference data from 1000 Genomes and the PAGE Study by super population. There are several unique descriptive characteristics of the genetic similarity distributions. The continental similarity of Native Hawaiian participants on principal components 1-3 primarily extends between European and East Asian reference samples. Latino participants have considerable genetic variation across all reference groups, with most participants showing similarity to European and American Indian reference samples.

**Figure 2:**
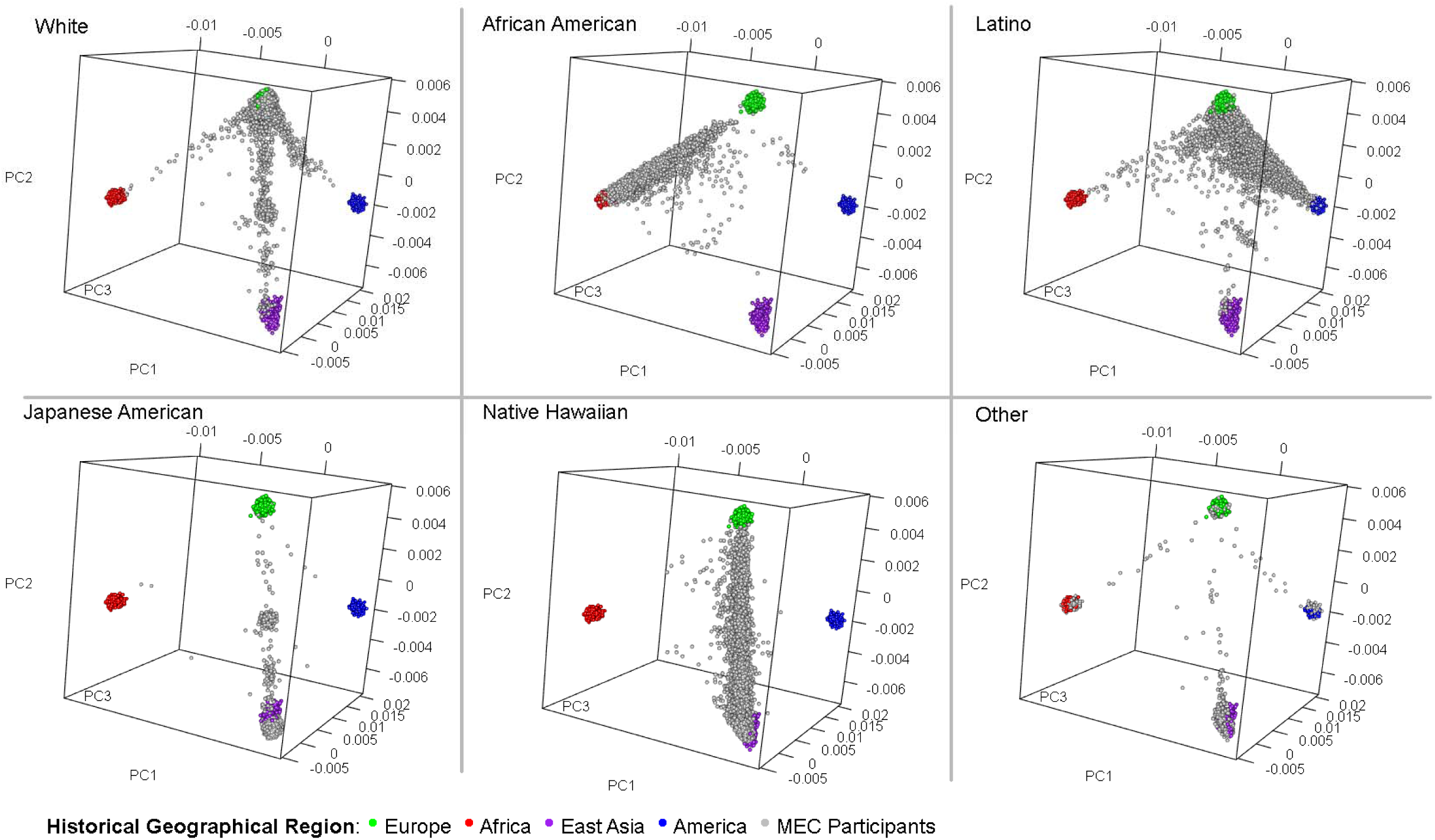
Genetics Database Principal Components. Principal components plotted in three dimensions (PCs 1,2,3) for each participant in the Genetics Database, separated by panel for each self-reported racial and ethnic group. Grey points are MEC participants and non-grey dots are participants from 1000G reference dataset and PAGE study.

Within each self-identified race and ethnicity, more detailed population characteristics can be identified across high-order principal components (supplemental figures 2-6). Notably, Mainland and Okinawan cluster groups can be identified within the Japanese American participants, along with first-generation admixed participants between the two groups (supplemental figure 5). Among Latinos, higher-order principal components illustrate considerable ancestral variation by geographical region across the Americas (supplemental figure 3). Reference data indicates Latino MEC participants’ genetic similarity aligns with reference data from South America, Mexico, and the Caribbean.

As an additional method to explore global similarity distributions, we estimated unsupervised clustering using ADMIXTURE with k=5 clusters (Figure 3). Within each self-reported racial and ethnic group, we observed a large variation in estimated cluster proportions, with many individuals across different self-reported race and ethnicity having similar values. While we choose to label clusters without consideration of self-reported participant characteristics within each cluster, a comparison of their distributions with reference data reveals that Cluster 1 reflects a high degree of African similarity, Cluster 2 aligns with Amerindigenous, Cluster 3 with East Asian, Cluster 4 with European, and Cluster 5 with Polynesian.

**Figure 3:**
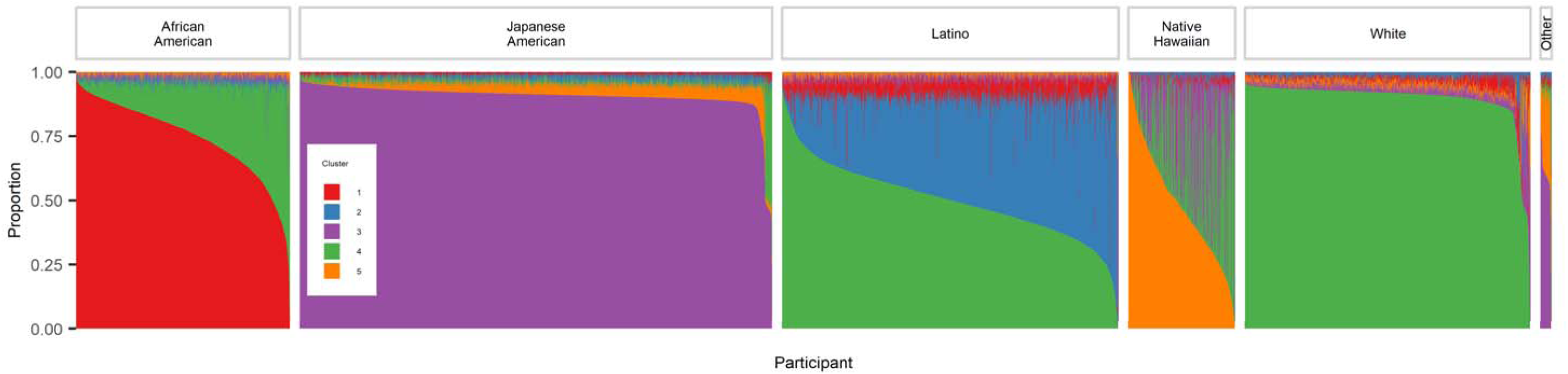
Clustered (k=5) Admixture Proportions. Unsupervised clustering of MEC Genetics Database participants using ADMIXTURE with k=5 clusters. The y-axis corresponds to portion of cluster, summing to 100% for each participant. The x-axis is each participant in the Genetics Database. Color-coded by cluster.

### Genetics Data Availability and Quality

Among variants with MAF > 1%, all racial and ethnic groups had a mean R^2^ >= 0.8, except for less common variants among Japanese American samples, indicating good imputation quality across all groups for common variants. The imputation quality of variants with MAF < 1% varied considerably across racial and ethnic groups, with the Japanese American and Native Hawaiian sub-samples having the poorest quality for rare variants, consistent with the under-representations of these populations in the imputation reference panel.^48,49^

### GWAS Results

To illustrate the use of these data in GWAS analyses, we performed association analyses for breast, colorectal, and prostate cancer in the Genetics Database with TOPMed imputed data. Variants with imputation quality R^2^ >= 0.8 (supplemental figure 7) were analyzed. We tested 5,117,063 variants in association with the three phenotypes and then filtered GWAS significant associations to variants lying outside of 500 kb around known regions for each phenotype to identify novel associations. Before filtering out known regions, we identified several significant GWAS associations for breast and prostate cancer in the admixed sample. Across GWAS, we observed minimal overdispersion (breast cancer λ = 1.03, colorectal cancer λ = 1.03, prostate cancer λ = 1.03; supplemental figure 8). After applying known region filter rules, no novel GWAS-significant variants remained, which is expected given the smaller relative case sample size within the MEC compared to the corresponding cancer GWAS consortiums^45–47^.

### Evaluation of Prior GWAS-identified Variants

In the multiethnic analyses of previously identified GWAS risk variants, 90/313 (28.8%) breast cancer variants, 61/205 (29.8%) colorectal cancer, and 183/450 (40.7%) prostate cancer variants were replicated at p ≤ 0.05 in the combined, multiethnic sample (supplemental figure 9, supplemental table 1). Ethnic-specific and multiethnic replication statistics are shown in supplemental tables 1-4. Two-hundred and forty-two (77.3%) breast cancer variants, 176 (85.9%) colorectal cancer, and 389 (86.4%) prostate cancer variants had a direction of effect consistent with the original discovery GWAS.

The inverse standard error weighted correlation between the log-odds (beta) estimates in the combined MEC sample and the analysis from estimates from prior GWAS-identified variants are R^2^=0.53 for prostate cancer, R^2^=0.18 for colon cancer, and R^2^=0.20 for breast cancer (supplemental figure 9). These correlations varied across self-reported race and ethnicity, with Latino and White samples having the strongest correlations. The mean effect sizes across phenotypes in the multiethnic sample were OR=1.05 for colorectal cancer and breast cancer and 1.06 for prostate cancer per risk allele. Given the discovery of these variants from large sample GWAS studies, these small allelic effect sizes are expected.

There was variation in heterogeneity of effect sizes by self-reported race and ethnicity. This heterogeneity was quantified using Cochrane’s Q and the Inconsistency Index, I^2^. The distributions of these statistics show 21/205 (10.2%) of colorectal cancer variants, 35/313 (11.2%) of breast cancer variants, and 69/450 (15.3%) of prostate cancer variants to have significant heterogeneity across racial and ethnic groups, defined by p-heterogeneity < 0.05. Prostate cancer had 240/450 (53.3%) variants with an I^2^ over 80%, colorectal cancer had 103/205 (50.2%), and breast cancer had 145/313 (46.3%).

### PRS Analysis

Evaluation of racial and ethnic-specific distributions of all weighted and non-standardized PRSs showed a lower mean PRS for breast cancer among White participants and a higher mean PRS for prostate cancer among African American participants. Hazard ratios and cumulative risk for breast cancer, colorectal cancer, and prostate cancer across race and ethnicity by PRS quantile are shown in Figure 4. The largest per-SD increase in risk was seen among White males for prostate cancer, where a 1-SD increase in PRS was associated with 2.13 (95%CI 1.93, 2.34) times the rate of prostate cancer. There was variation in the effect size and standard error between PRSs and each cancer type across racial and ethnic groups. This variation in SEs across racial and ethnic groups originates not only from differences in person-years and case counts but also PRS performance (as seen in the PRS variant replication). The absolute risk (cumulative and 5-year) of prostate cancer and breast cancer was considerably higher than that of colorectal cancer reflecting both higher baseline risk and PRS effects. At age 80, a self-reported African American male at 95% genetic risk percentile has a 43.1% (36.9%, 50.9%) lifetime risk of prostate cancer, whereas a White male at the same age and PRS has a risk of 35.9% (29.9%, 43.8%) (Figure 4, Supplemental Table 5). On the 5-year risk scale, the African American male’s 5-year risk would be 0.6% (0.5%, 0.6%), and the White male’s risk would be 0.9% (0.8%, 1.0%) (Supplemental Figure 11). The lower 5-year risk reflects the effect of competing risks affecting risk over the next 5 years.

**Figure 4:**
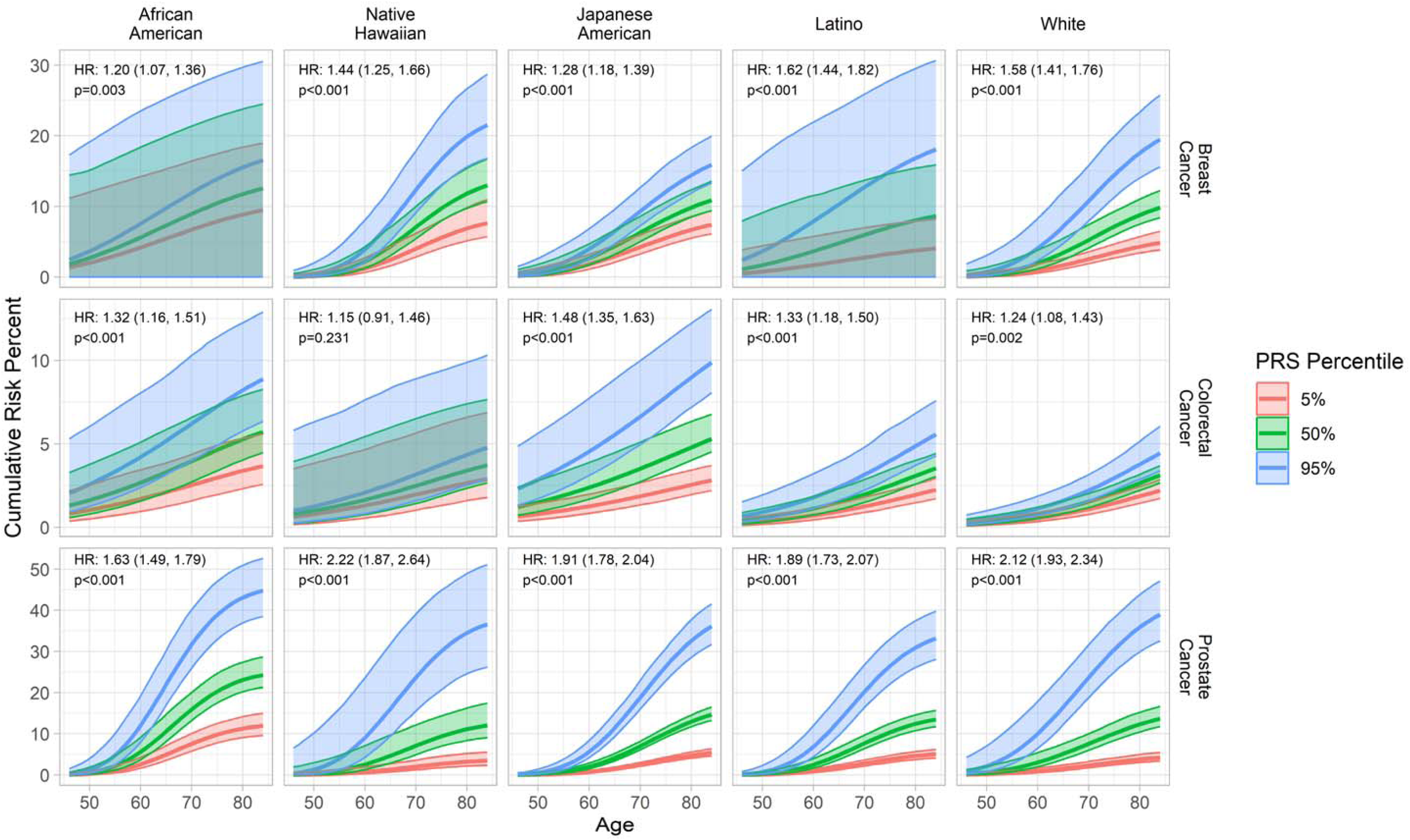
Absolute Risk Curves for Breast Cancer, Colorectal Cancer, and Prostate Cancer. Hazard ratios (per standard deviation increase) and cumulative risk of Breast, Colorectal, and Prostate cancer stratified by self-reported racial and ethnic group color-coded by PRS quantile, within the Genetics Cohort.

### Correlations between Anthropometric Measures, Diet Quality, Lifestyle Factors, Comorbidities, Population Descriptors, and Genetic Similarity Measures

We conducted a series of correlation analyses to illustrate the association between genetic similarity (i.e. principal components) and race and ethnicity across a collection of self-reported baseline variables and cancer (supplemental figures 12-15). These results are presented in categories of anthropometric measures, diet quality, lifestyle factors, and cancer. Anthropometric measures had the strongest correlations with genetic and non-genetic population descriptors. For example, principal components explained 30% of the variation in height, and self-reported race and ethnicity explained 27% (supplemental figure 15). When both race and ethnicity and principal components are modeled jointly, both variables explain 30% of the variation in height, illustrating a similar predictive ability of both variables for this phenotype. The 3% difference in predictive ability between these two measures is evident in the ethnic-specific correlation analyses. For example, when restricting the sample to Native Hawaiian participants, principal components still capture variation in height, explaining 11% of the variation within the sub-sample. For lifestyle risk factors, we still observe considerable variation explained by race and ethnicity or principal components indicating genetic and non-genetic descriptors are associated with additional unmeasured variables that determine or are correlated with nSES. This analysis highlights the need for careful consideration of the use of population descriptors in risk analysis as these variables often are highly correlated with other risk factors. Following recent National Academy of Sciences, Engineering, and Medicine (NASEM) recommendations, researchers should directly evaluate these risk factors when available.

### Absolute Risk by nSES and PRS

Using joint models to understand the independent effects of each variable on disease risk, we modeled cancer as an effect of PRS and nSES stratified by race and ethnicity. Using these models, we estimated cumulative risk by age (supplemental figure 16, 17). The PRS risk stratification we observed in this analysis was similar to the prior PRS models.^45–47^ However, we now observed additional, minor risk variation by nSES within the PRS percentile. For example, among African American males who were at the 95% PRS risk quantile, those in the 0%-25% nSES group had a 49.8% chance of developing prostate cancer by age 85, whereas those in the 76%-100% nSES group had a 57.5% chance of developing prostate cancer at the same age (supplemental figure 17). We illustrate the added value of each variable in risk estimation in Table 2 for four hypothetical individuals with a known nSES and PRS. Individual 1 is an African American male with a PRS at the 95^th^ percentile who is at the lowest quartile of nSES. When estimating cumulative risk at age 85 using only age and race and ethnicity, we see a cumulative risk of 26.8%. This is the same risk as individual 2, who has a different PRS and nSES. However, since PRS and nSES are associated with prostate cancer risk, each additional variable adds value to risk stratification. nSES explains minimal variation in prostate cancer risk, resulting in a small change in estimated risk when this variable is included (26.8% versus 25.6%). When PRS is included in the model, individual 1’s risk moves from 27.8% to 45%. When all variables are included in risk prediction, this individual has an estimated 49.8% chance of having had prostate cancer by age 85, versus the 27.8% using only age and self-reported race and ethnicity.

**Table 2:** Predicted Cumulative Risk of Prostate Cancer at age 85, given predictor variables. Predicted cumulative risk of prostate cancer at age 85 for four hypothetical individuals. Predicted cumulative risk shown using for models including different sets of predictor variables. The base model is age + race and ethnicity. The second model adds PRS to the base model, the third model adds nSES to the base model, and the final model adds both PRS and nSES to the base model.

|  | Predicted Cumulative Risk of Prostate Cancer at age 85, given prediction variables: |  |  |  |
| --- | --- | --- | --- | --- |
|  | Age + race and ethnicity | +PRS | +nSES | +PRS +nSES |
| Individual 1 (race/ethnicity = African American, PRS = 95%, nSES = Q1) | 26.77% | 45.00% | 25.64% | 49.79% |
| Individual 2 (race/ethnicity = African American, PRS = 5%, nSES = Q4) | 26.77% | 12.02% | 30.96% | 10.50% |
| Individual 3 (race/ethnicity = White, PRS = 95%, nSES = Q1) | 16.52% | 39.56% | 14.63% | 31.62% |
| Individual 4 (race/ethnicity = White, PRS = 5%, nSES = Q4) | 16.52% | 4.29% | 17.82% | 5.66% |

### Assessment of genetic risk in the combined MEC samples

Using the same modeling techniques as above, we estimated the cumulative risk of prostate cancer in a multiethnic model to compare cumulative risk estimates between self-reported race and ethnicity stratified models and the genetic similarity model. In this analysis, cancer risk is modeled as a function of PRS and the population descriptor of interest. We fit two models to compare estimates: one using the population descriptor self-reported race and ethnicity and the second using genetic similarity cluster variables (k=5).

In the self-reported race and ethnicity model, an individual who self-reports as an African American male at the 90% PRS percentile would be estimated to have a prostate cancer cumulative risk of 17.1% at age 85 (supplemental figures 18, 19). If replacing self-reported race and ethnicity with a measure of genetic similarity (k=5) in a multiethnic model, an individual with a genetic similarity characteristic of an average African American participant (cluster 1=85% and cluster 2=15%) and the same PRS of 90%, would have a similar cumulative risk of 16.2%.

Across most cancer types and self-reported racial and ethnic groups, we observed similar performance between a self-report and genetic similarity approach. In populations with greater heterogeneity of genetic similarity (e.g., Native Hawaiians), risk estimation at genetic similarity estimates (clusters) that more closely align with the observed distribution in the population produce cumulative risk estimates most similar to a self-reported model.

## Discussion

The lack of diversity in genomics data is a critical issue discussed by the scientific community, as it may lead to the exacerbation of health disparities and hinder the translational impact of genetic findings^23–28^. Resolving this issue has been the goal of several national initiatives.^50–52^ Although the disparity in ancestral representation has been shrinking, many currently available data sources are limited by their degree of diversity, study design, and availability of non-genomic covariates. Extensive genetic diversity across populations remains unexplored, especially within a single, prospective cohort. The MEC Genetics Database and Genetics Cohort provide a unique opportunity to explore potentially overlooked important markers of cancer susceptibility and facilitate the development of risk models accounting for population-specific allelic heterogeneity.^24–27^

### Descriptive and Analytical Characteristics

Our descriptive and analytical results highlight the unique and valuable characteristics of the MEC Genetics Database and the nested Genetics Cohort consisting of incident cases. We observed high genetic diversity across many populations, representing genetic variation within the five populations initially targeted for recruitment. Due to the nested nature of these data, both the Genetics Database and Genetics Cohort contain detailed baseline and follow-up information on participants’ lifestyle, diet, comorbidity, reproductive, and environmental factors collected across follow-up. We found few differences between the complete MEC, the MEC Genetics Database, and the MEC Genetics Cohort across numerous covariates’ means and prevalence measures. This characteristic supports the generalizability of findings across data sources within the MEC. Regarding the analysis, the GWAS and replication results support the validity of the data source and analytical methods. Given our model specifications, we did not identify any overdispersion issues, and we observed an expected distribution of replication effect sizes in the combined multi-ancestry sample. These results illustrate comprehensive control of population stratification, cryptic relatedness, and batch/project effects that may be identified through overdispersion. When used in trans-ancestry meta-analyses and fine-mapping, the MEC Genetics Database is, and will continue to be, a resource for samples of admixed ancestry, contributing to the discovery and characterization of risk variants for cancer and other chronic diseases.^31^

In the replication phase of this study, we found significant heterogeneity for 21 out of 205 colorectal cancer variants, 35 out of 313 breast cancer variants, and 69 out of 450 prostate cancer variants tested across all racial and ethnic groups. Research supports the idea of homogeneous SNP effects across populations.^53–55^ The heterogeneity we observe may be due to differing LD structures between populations, resulting in tagging SNPs that ineffectively capture genetic variation associated with the phenotypes. Alternatively, heterogeneous effects may also be observed through epistasis and/or gene-environment interactions, where the prevalence of the interacting component (allele frequency or prevalence of the lifestyle/environmental component) varies across populations, resulting in different observed effect sizes.^56^ Given the vast information on non-genetic risk factors, including environmental exposures (e.g. air pollution), the MEC has the information potentially to address these questions.

### Genetic Risk Score Analyses

In addition to discovery, the MEC Genetics Database is a powerful resource for PRS evaluation and characterization, where subsets of this data have already been used for these analyses.^29,30^ Individual-level time to event data has allowed us to estimate absolute risk within levels of several variables. Current methods for estimating absolute risk, such as iCare, use external cancer incidence and all-cause mortality summary statistics to estimate absolute risk across and within populations.^31,57^ This is done since most biobanks have complex, often hospital-based, sampling that does not allow for valid baseline risk estimation within the sample. In contrast to using external baseline risk information, we tabulated time-to-event data into lifetables across self-reported race and ethnicity, sex, PRS quantile, and ancestry quantile, then used this tabulated data to calculate model-based and empirical absolute risk across covariate levels. This stratified analysis allows for baseline risk and cumulative survival probabilities to vary across covariate levels. This is important in cases of pleiotropic traits (i.e. traits that are associated with variants that affect the risk of competing events). Thus, we are not required to assume that baseline and age-specific risk is the same across exposure and covariate levels within and between race, ethnicity and ancestry.

In our PRS analyses, we estimated hazard ratios, incidence, cumulative risk, and 5-year risk by PRS level. Although we observed variation in the effect sizes and performance of PRS across racial and ethnic groups, the absolute risk, in many cases, was higher among racial and ethnic groups with smaller effect sizes. This pattern is seen most clearly with PRS performance across prostate cancer. The random error in these estimates was estimated using standard error from model variable estimates; however it is important to note that there is additional random error in an individual’s estimated PRS value itself that will further affect uncertainty of the cumulative risk estimates.^58^ To ensure the accuracy of our baseline risk, age effects, and survival estimation, we compared the 50% PRS risk group (i.e., average risk) against SEER age-specific cumulative risk estimates and incidence. This comparison showed close agreement in estimates of age-specific cumulative risk and incidence.^4^

### Interaction Analyses and Population Descriptors in Genetic Risk Estimation

We illustrate the complex and often strong associations between genetic similarity, self-reported race and ethnicity, baseline characteristics, and incident cancer in several analyses. In the context of PRS effect estimation and risk prediction, the relationship between these variables makes it difficult to parse the individual variable effects and estimate strata-specific cumulative risk.^59^ Recent reports, such as the National Academy of Sciences, Engineering, and Medicine (NASEM) report “Using Population Descriptors in Genetics and Genomics Research,” and later commentaries, such as the “Guidance on Use of Race, Ethnicity, and Geographic Origin as Proxies for Genetic Ancestry Groups in Biomedical Publications,” discuss many of these complexities and possible pitfalls, misuse, and misinterpretation of results using population descriptors and genetic similarity^19,60^. The relationship between population descriptors, common genetic variation, and non-genetic covariates have been established in separate datasets; however, in our analyses, we illustrate these associations in a single dataset and in joint models.

Genetic similarity measures such as principal components and ancestry clusters, often interpreted as “ancestry percentage,” are commonly used in genetic epidemiology. These variables are sometimes used as indicators of genetic risk since they are estimated using genetic variation. Our correlation analysis illustrates how this form of interpretation is often invalid. We illustrate the strong association between population descriptors (i.e., race and ethnicity) and these measures of genetic similarity, showing that these genetic similarity variables are indicators of other population descriptors (e.g., race and ethnicity), which are in turn strongly associated with non-genetic exposures (e.g., smoking prevalence).

Within self-reported race and ethnicity for many variables, we still observe phenotypic variation explained by principal components in the correlation analysis. Although this stratified analysis restricts each race and ethnicity to remove most variation explained by principal components, residual variation in principal components may still capture variation in non-genetic risk factors. This phenomenon can best be observed in “control” variables that are not expected to be directly caused by common genetic variation.

### Comparison of Risk Estimation using a Self-report and Genetic Similarity Approach

Population descriptors provide insight into health disparities. A comparison of these risk estimates across models illustrates how measures of disease risk by continental genetic similarity are essentially the same as using self-reported race and ethnicity, likely reflecting the strong association between these genetic similarity values and self-reported race and ethnicity, which also may be associated with common non-genetic risk factors. These PRS analyses highlight the point in the NASEM report’s guidance to “replace or supplement descent-associated population descriptors with information about the relevant factors that mediate differences in environmental exposures.” In models using population descriptors to generate risk estimates, individuals within groups represent the average covariate distribution within that descriptor level. In addition, the use of genetic similarity may not accurately capture or align with the underlying risk factors. Only inclusion of the actual disease risk factors in the model may represent the true tapestry of risk. Thus, a diverse and admixed cohort with varied environmental and lifestyle levels, such as the MEC, allows for examination of these effects in joint models. A model type that does not rely on self-reported race and ethnicity is flexible in that genetic similarity values are continuous and can be modeled jointly with genetic and non-genetic risk factors that may be associated with population descriptors.

## Conclusion

Identifying and testing genetic markers of cancer susceptibility across admixed racial and ethnic populations holds immense potential with widespread applicability and significance. Inclusion of admixed populations in genetics research can provide a clearer understanding of the spectrum of allelic variation that underlies population risk and disease heritability, facilitate the development of risk models accounting for allelic heterogeneity between populations, and contribute to the formulation of preventive strategies targeting individuals at high risk. The comprehensive nature of these data provides opportunities for improved fine-mapping analyses, cross-population variant and PRS replication, absolute risk estimation, and gene-environment interaction analyses.

Our presented results show the extensive genetic diversity and comprehensive participant data as a rich resource for future genetic and epidemiological research. Future research using our Genetics Cohort can include more nuanced analyses such as fine mapping, cross-population variant replication, and gene-environment and gene-lifestyle interactions exploration. Such research is vital for addressing the critical issue of diversity in genomics and reducing health disparities. Additionally, the database’s genetic diversity highlights its value in assessing progress toward equitable, personalized medicine. Overall, the MEC Genetics Database sets the stage for groundbreaking work in understanding complex genetic interactions and their implications for public health and personalized healthcare across diverse populations.

## Supporting information

Supplemental Table 2 - replication_results_1kp3_imputed_BC

Supplemental Table 4 - replication_results_1kp3_imputed_PC

Supplemental Table 3 - replication_results_1kp3_imputed_CRC

## Data Availability

All data produced in the present study are available upon reasonable request to the Multiethnic Cohort Study (https://www.uhcancercenter.org/for-researchers/mec-data-sharing) and The database of Genotypes and Phenotypes (dbGaP).

https://www.ncbi.nlm.nih.gov/projects/gap/cgi-bin/study.cgi?study_id=phs002183.v2.p1

## Supplemental Materials

### Supplemental Figures

**Supplemental Figure 1:**
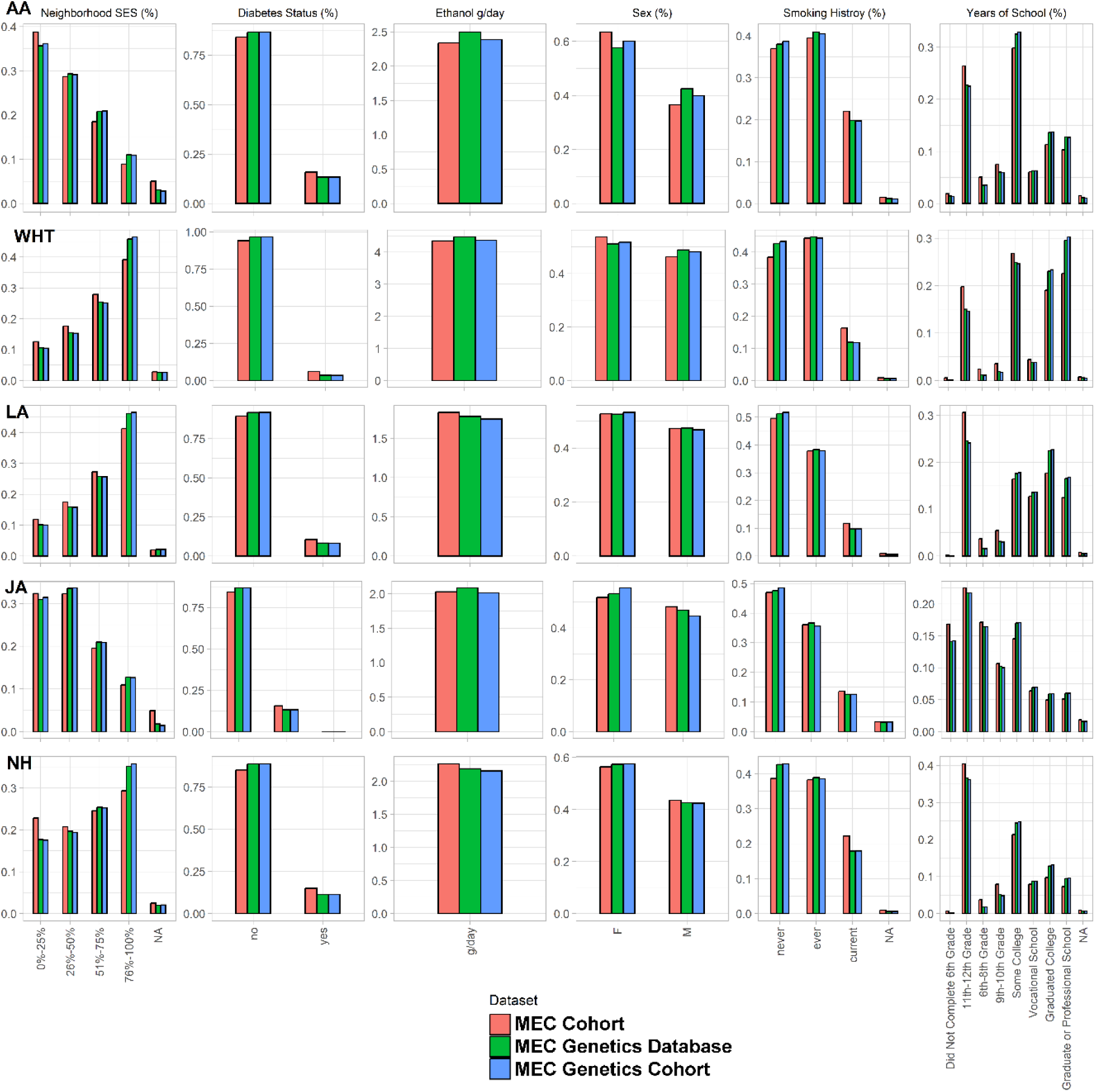
Participant Characteristics Across Samples. AA = African American, WHT = White, LA = Latino, JA = Japanese American, NH = Native Hawaiian

**Supplemental Figure 2:**
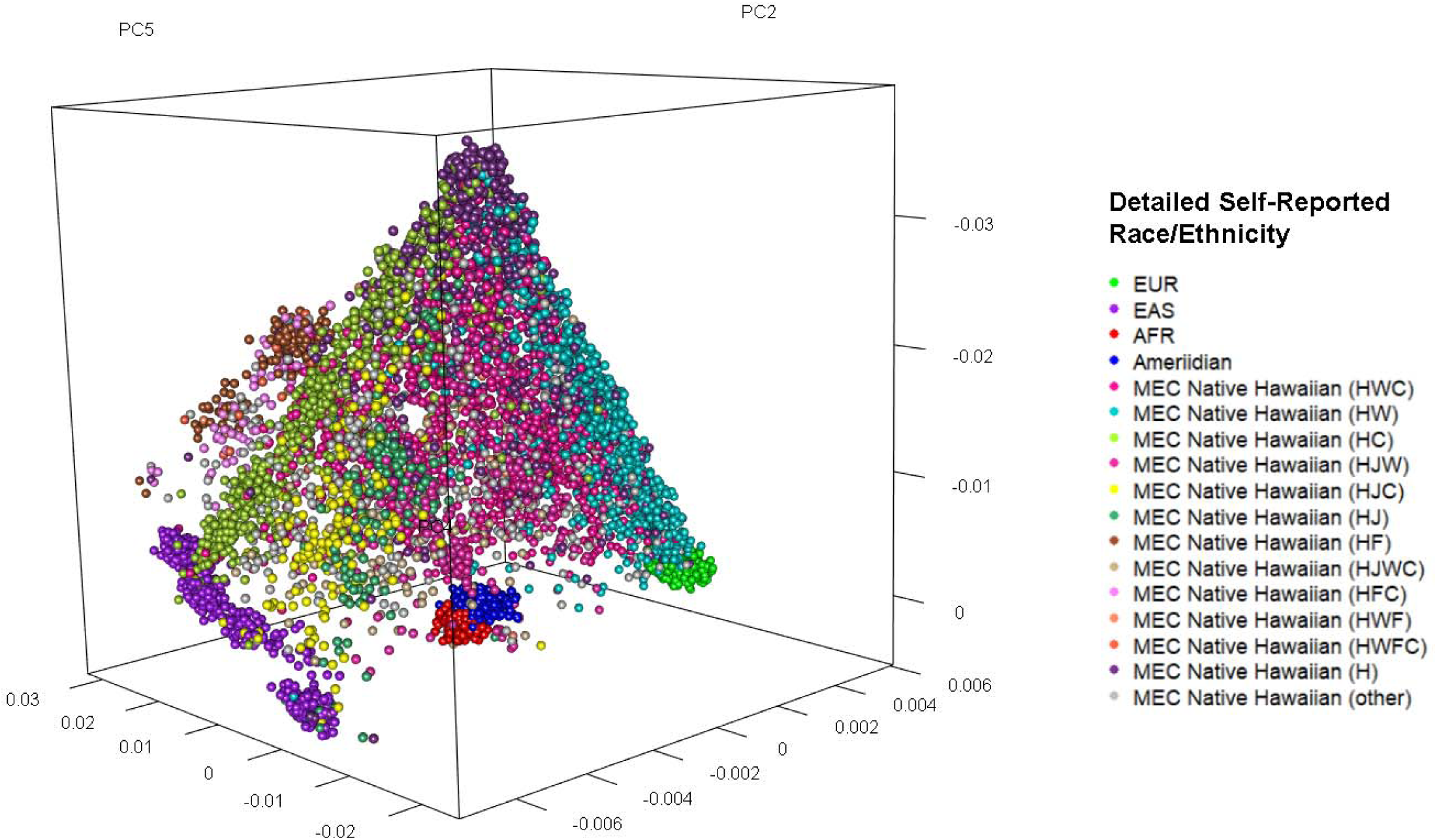
PC2xPC4xPC5 for MEC Native Hawaiians colored by composite self-report variable

**Supplemental Figure 3:**
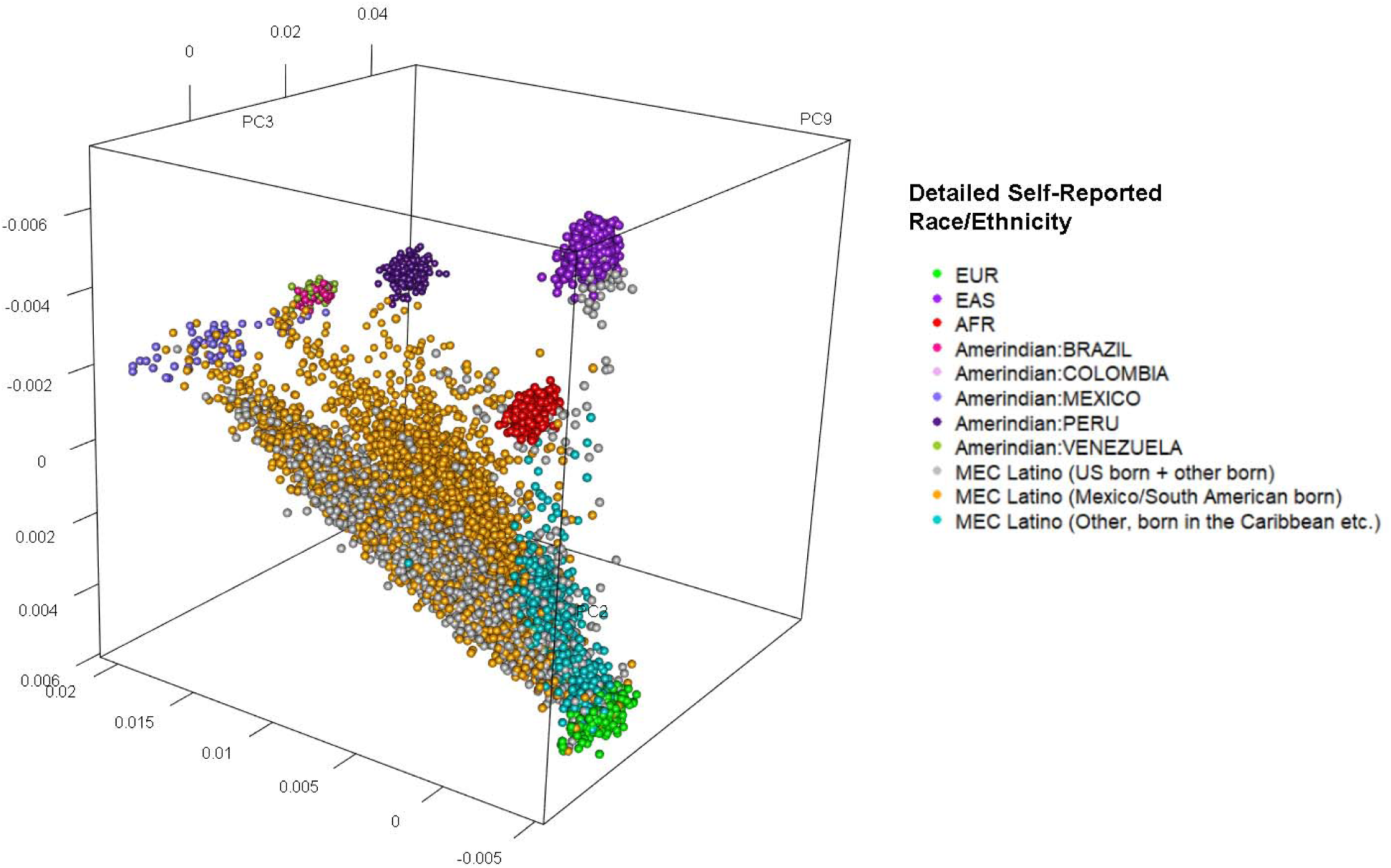
PC2xPC3xPC9 for MEC Latinos colored by birth area from baseline questionnaire

**Supplemental Figure 4:**
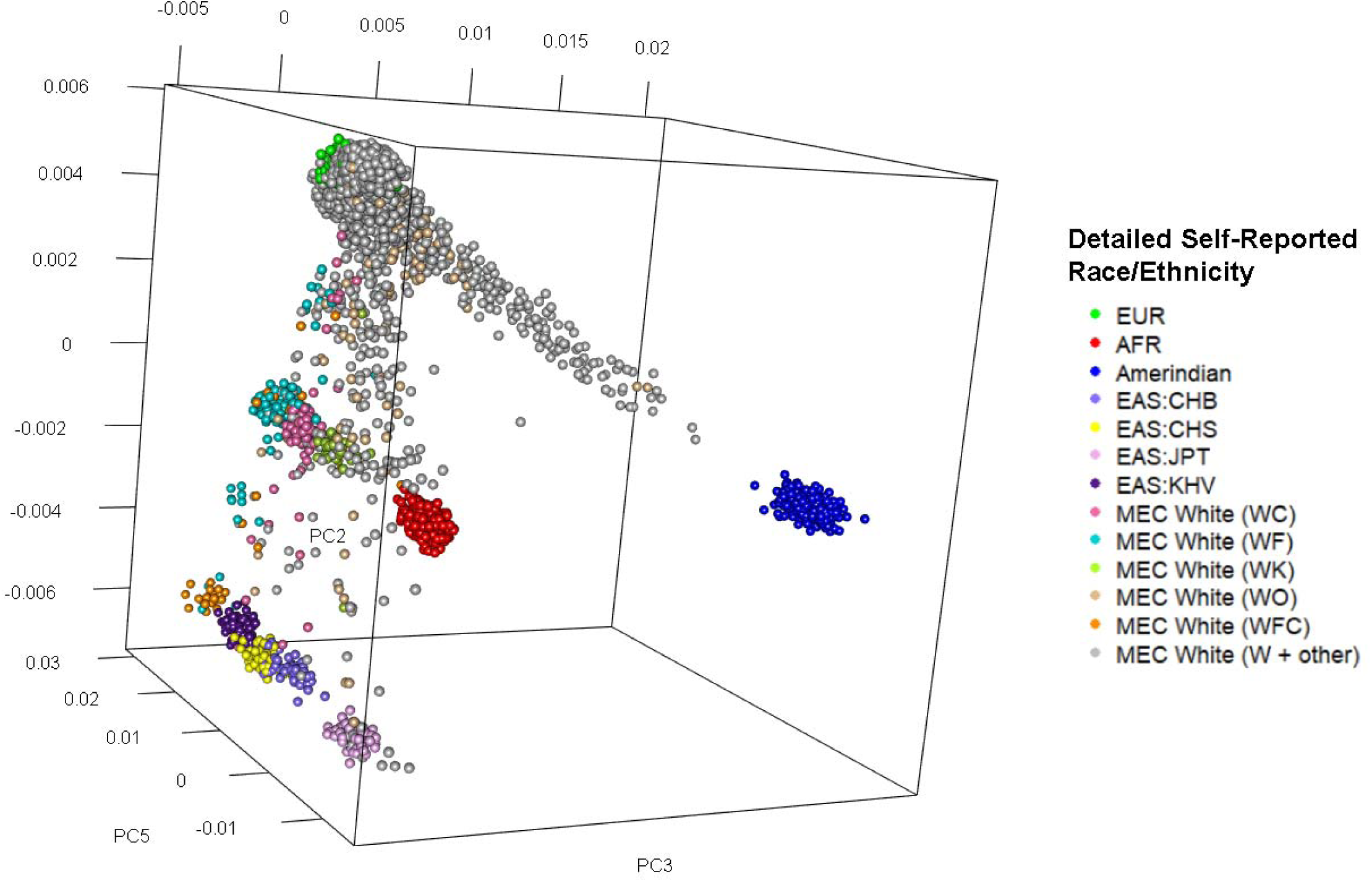
PC2xPC3xPC5 for MEC Whites colored by composite self-report variable

**Supplemental Figure 5:**
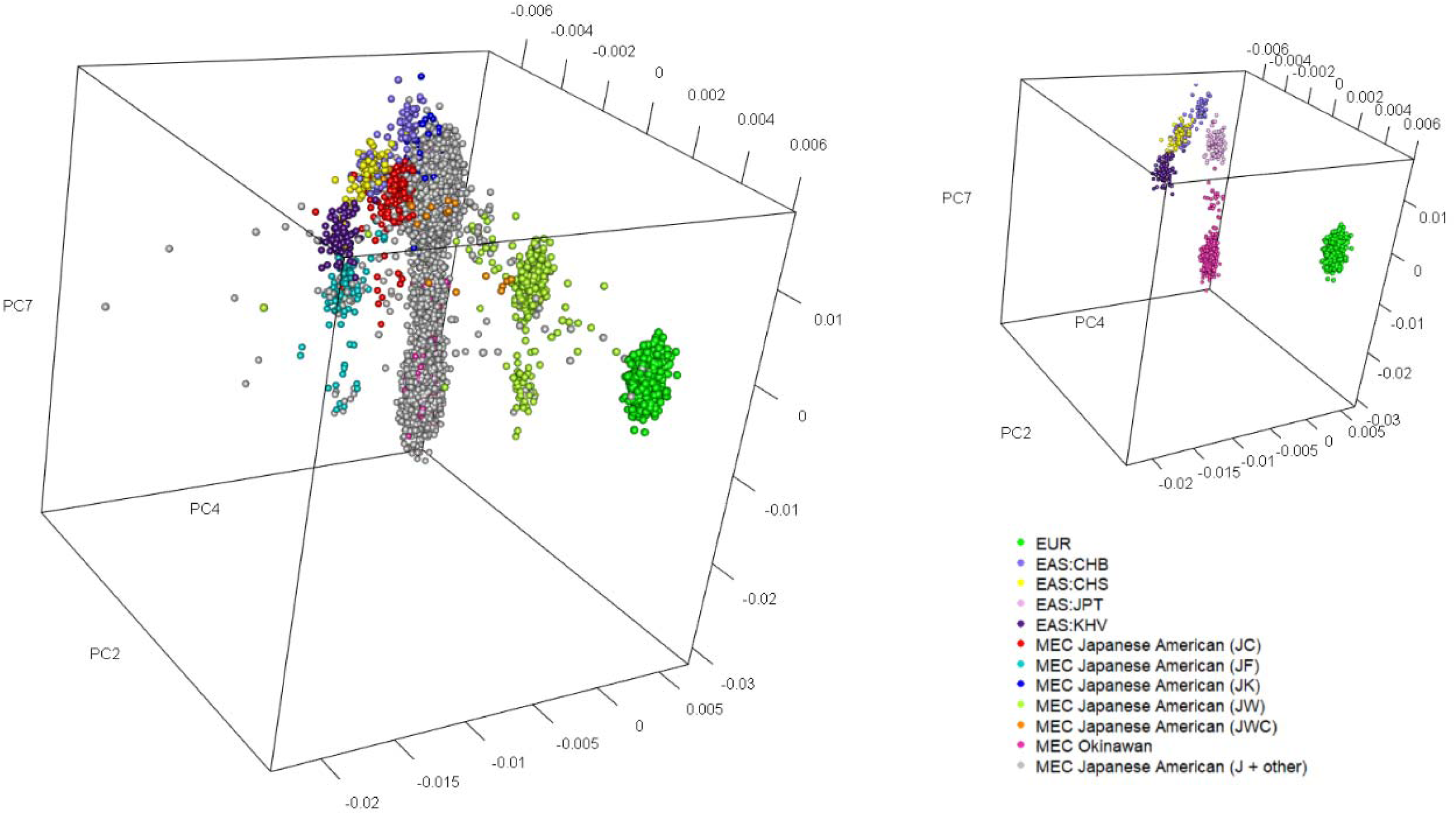
PC2xPC4xPC7 for MEC Japanese Americans colored by composite self-report variable and MEC Okinawan group. Top right small figure shows all the reference subjects and MEC Okinawan group only

**Supplemental Figure 6:**
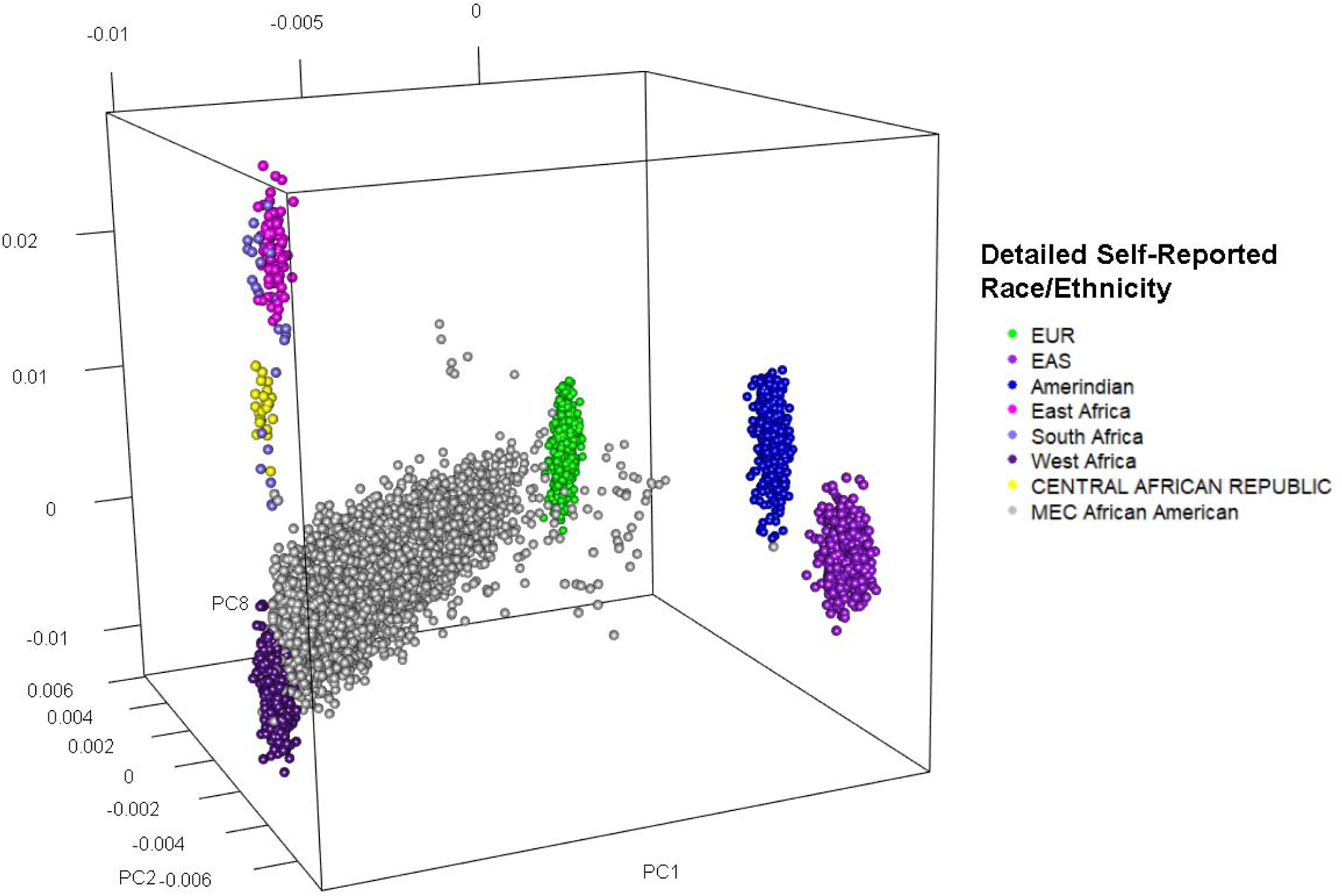
PC1xPC2xPC8 for MEC African Americans where African reference subjects were grouped into larger regions

**Supplemental Figure 7:**
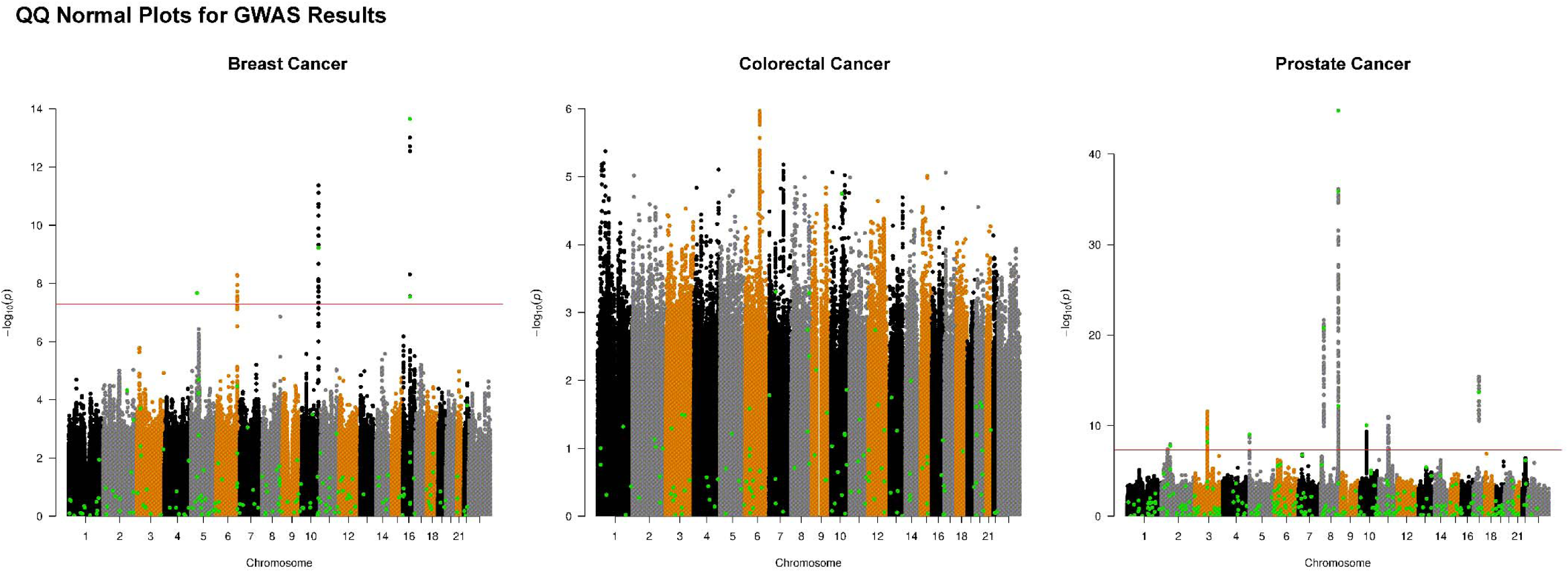
GWAS Manhattan Plots for Breast Cancer, Colorectal Cancer, and Prostate Cancer

**Supplemental Figure 8:**
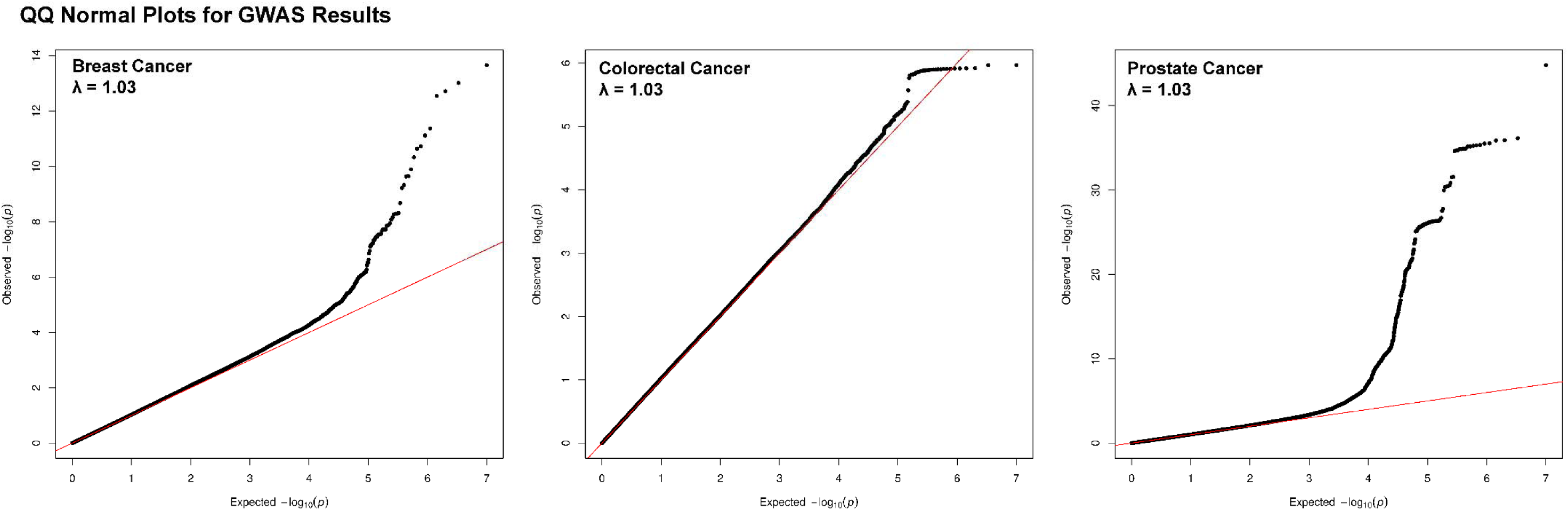
GWAS QQ Plots for Breast, Colorectal, and Prostate Cancer

**Supplemental Figure 9:**
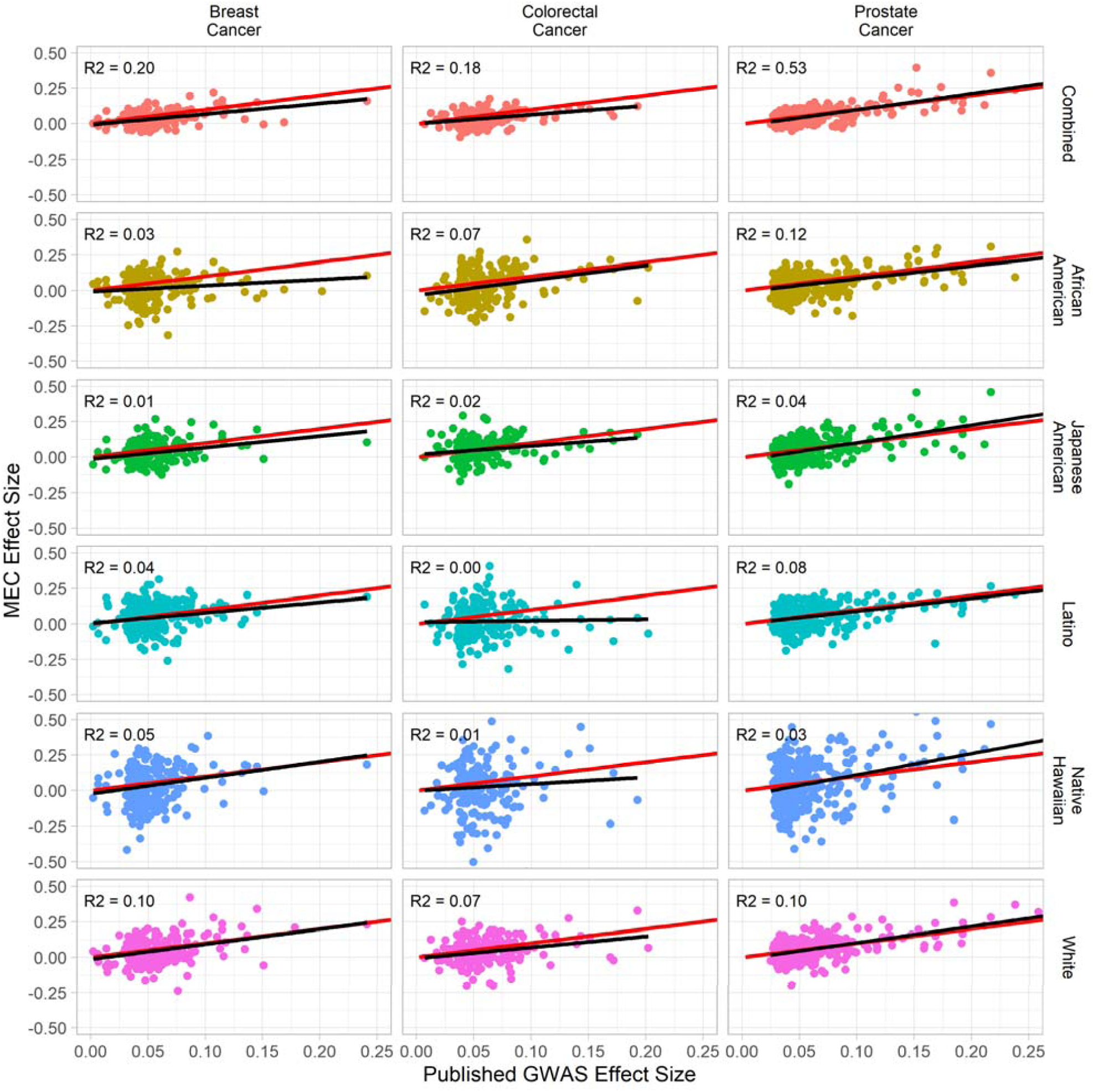
Replication analysis beta values plotted against external GWAS effect sizes by phenotype and race and ethnicity

**Supplemental Figure 10:**
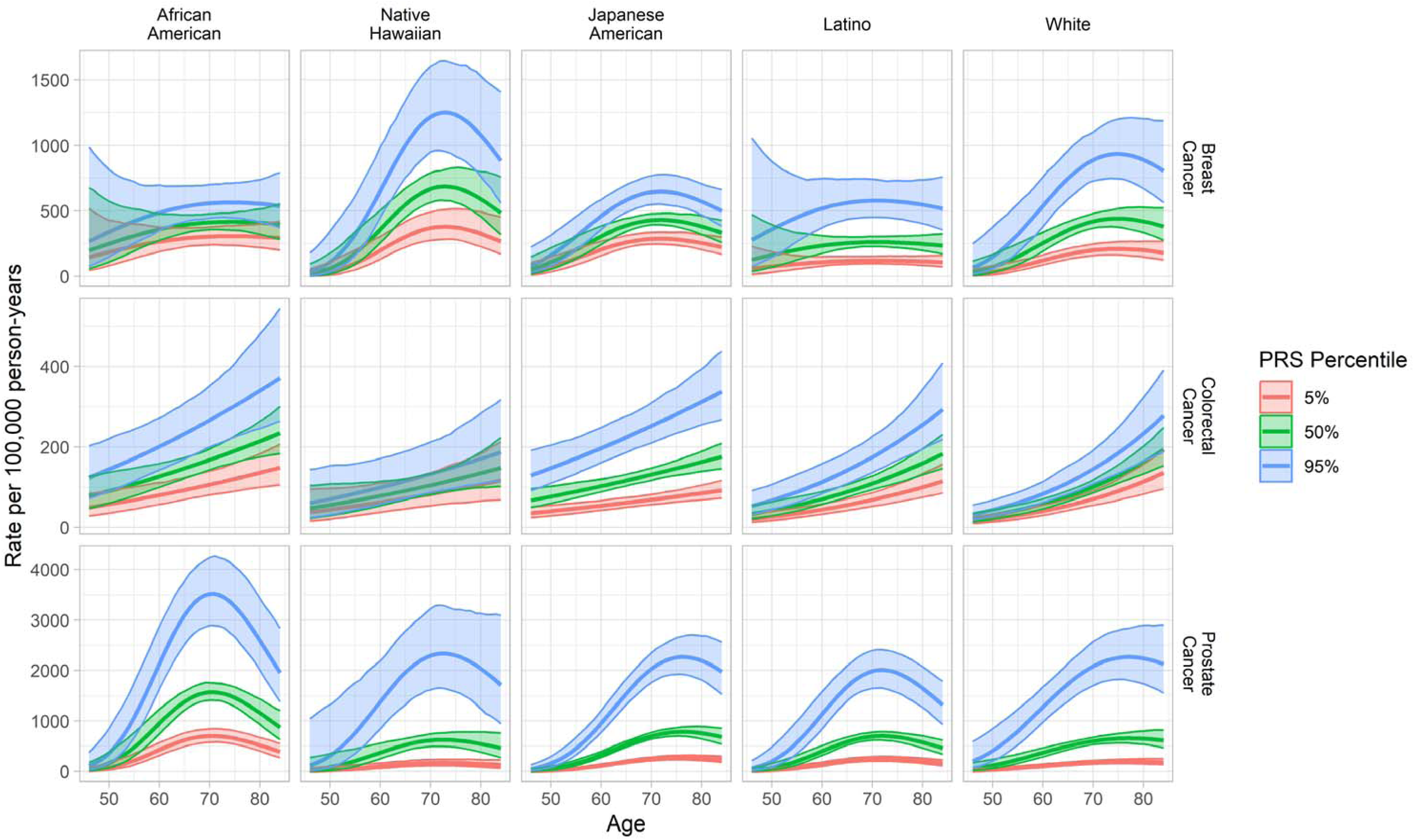
Incidence rates of BC, CRC, and PC by race and ethnicity and PRS quantile

**Supplemental Figure 11:**
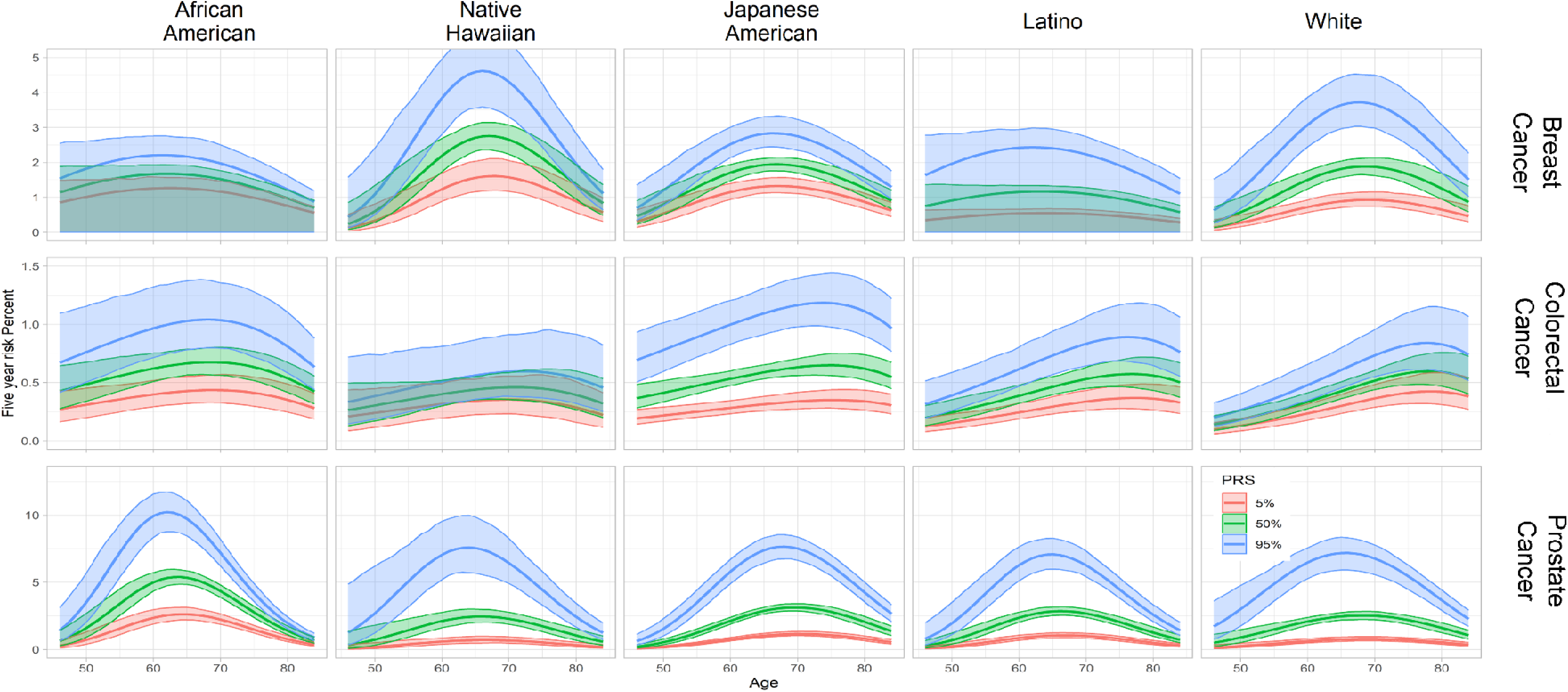
Five-year risk of BC, CRC, and PC by race and ethnicity and PRS quantile

**Supplemental Figure 12:**
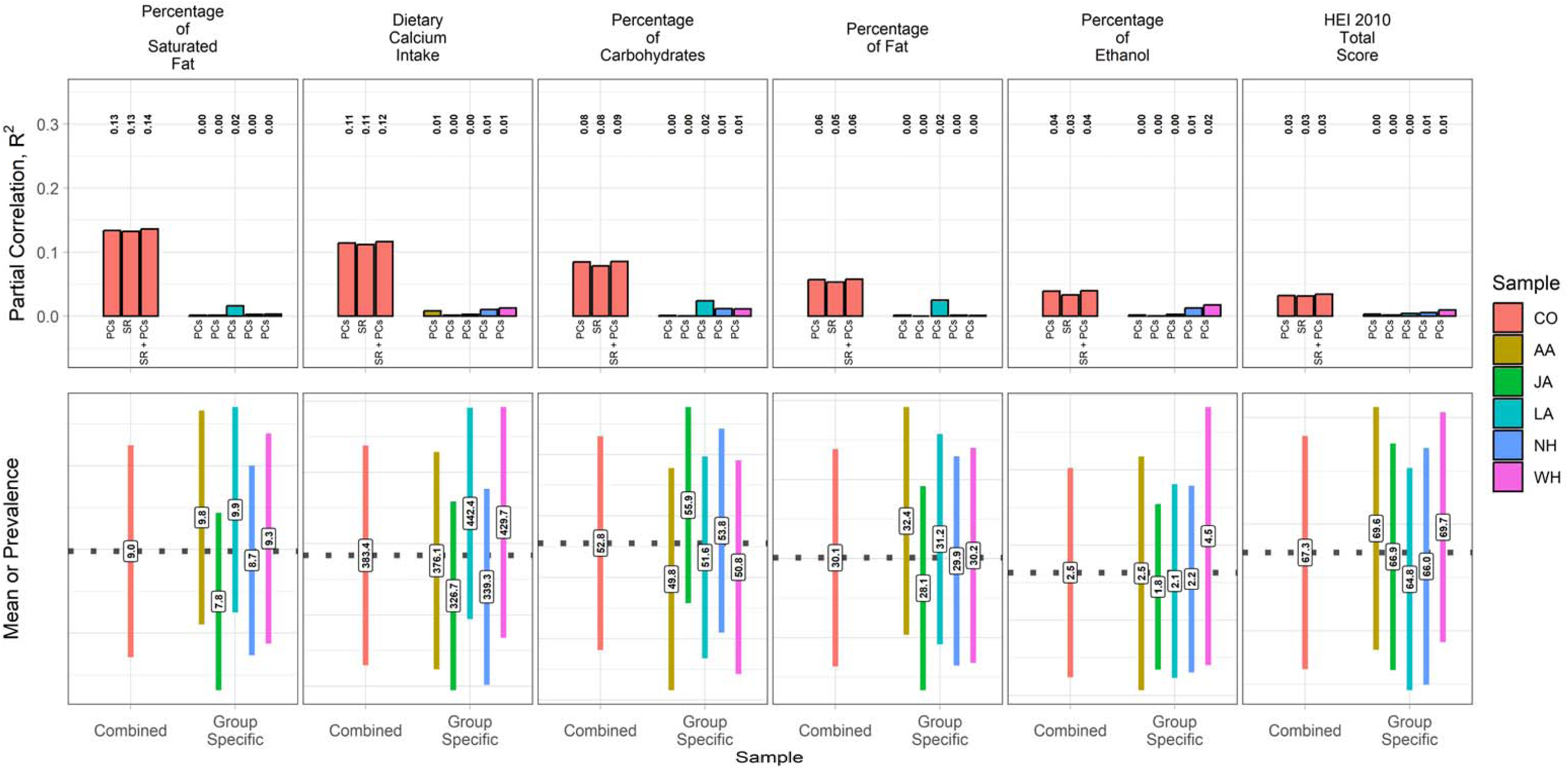
Partial Correlations of Dietary Factors. CO = Combined, AA = African American, JA = Japanese American, LA = Latino, NH = Native Hawaiian, WH = White

**Supplemental Figure 13:**
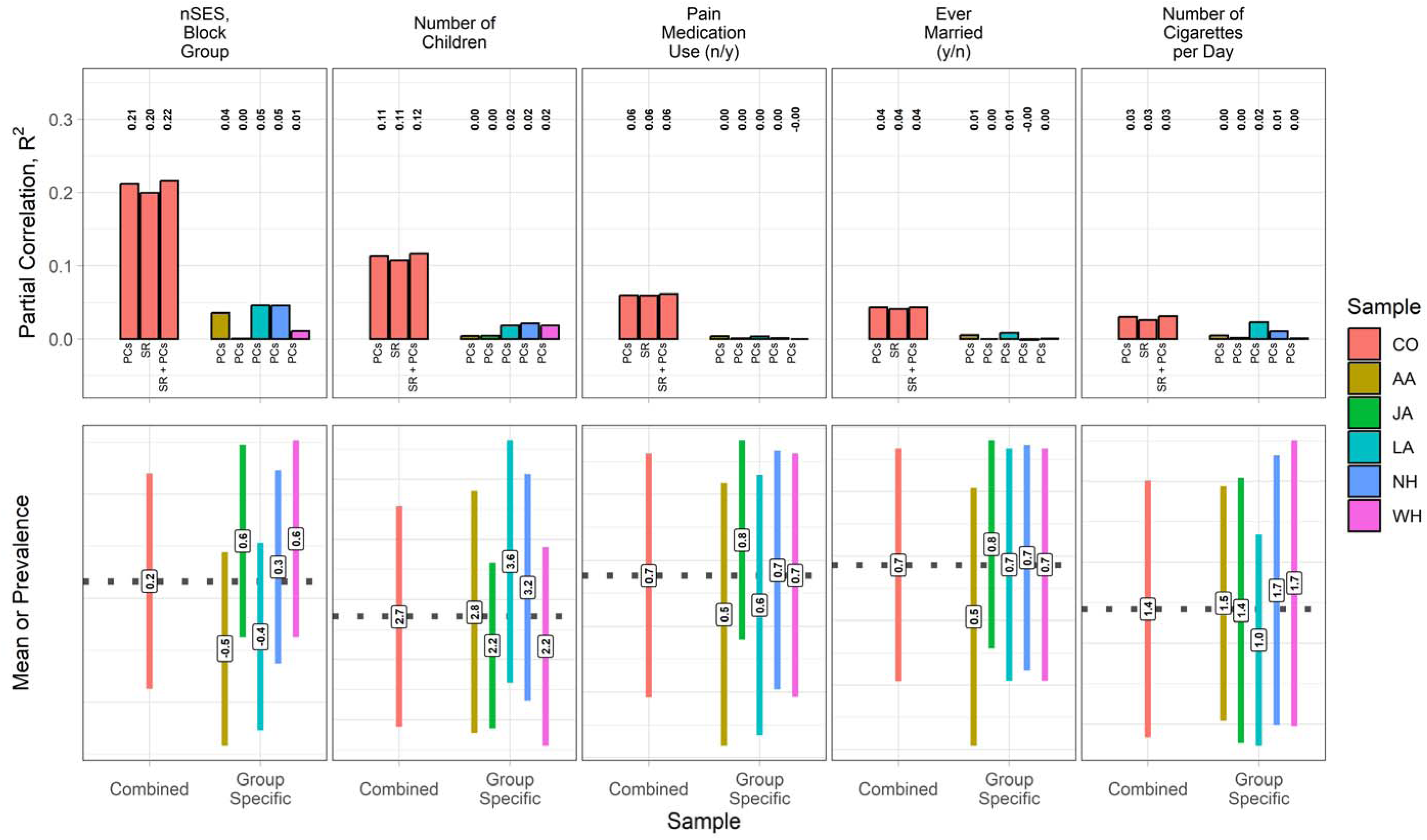
Partial Correlations of Lifestyle Factors. CO = Combined, AA = African American, JA = Japanese American, LA = Latino, NH = Native Hawaiian, WH = White

**Supplemental Figure 14:**
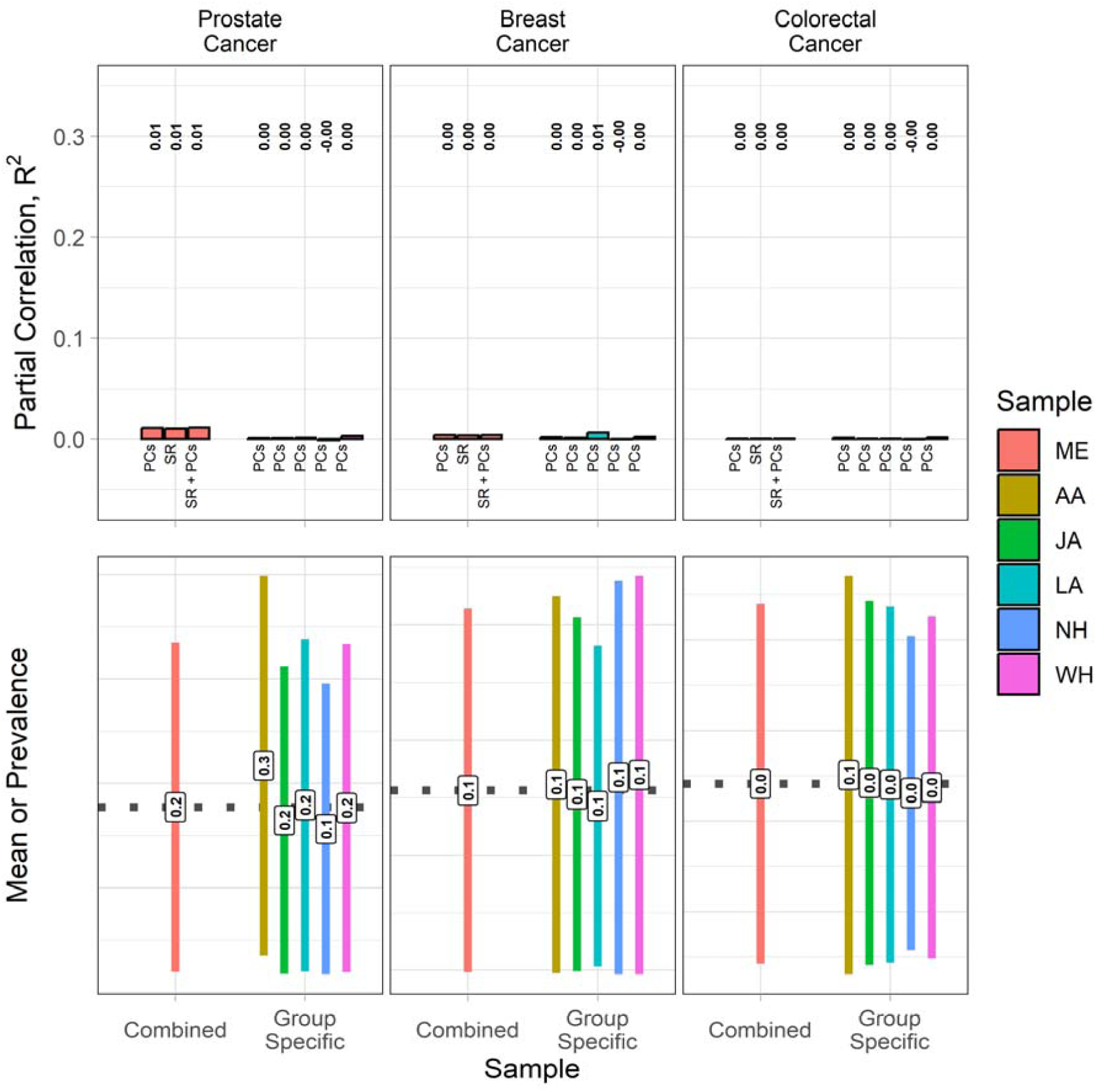
Partial Correlations of Cancer. CO = Combined, AA = African American, JA = Japanese American, LA = Latino, NH = Native Hawaiian, WH = White

**Supplemental Figure 15:**
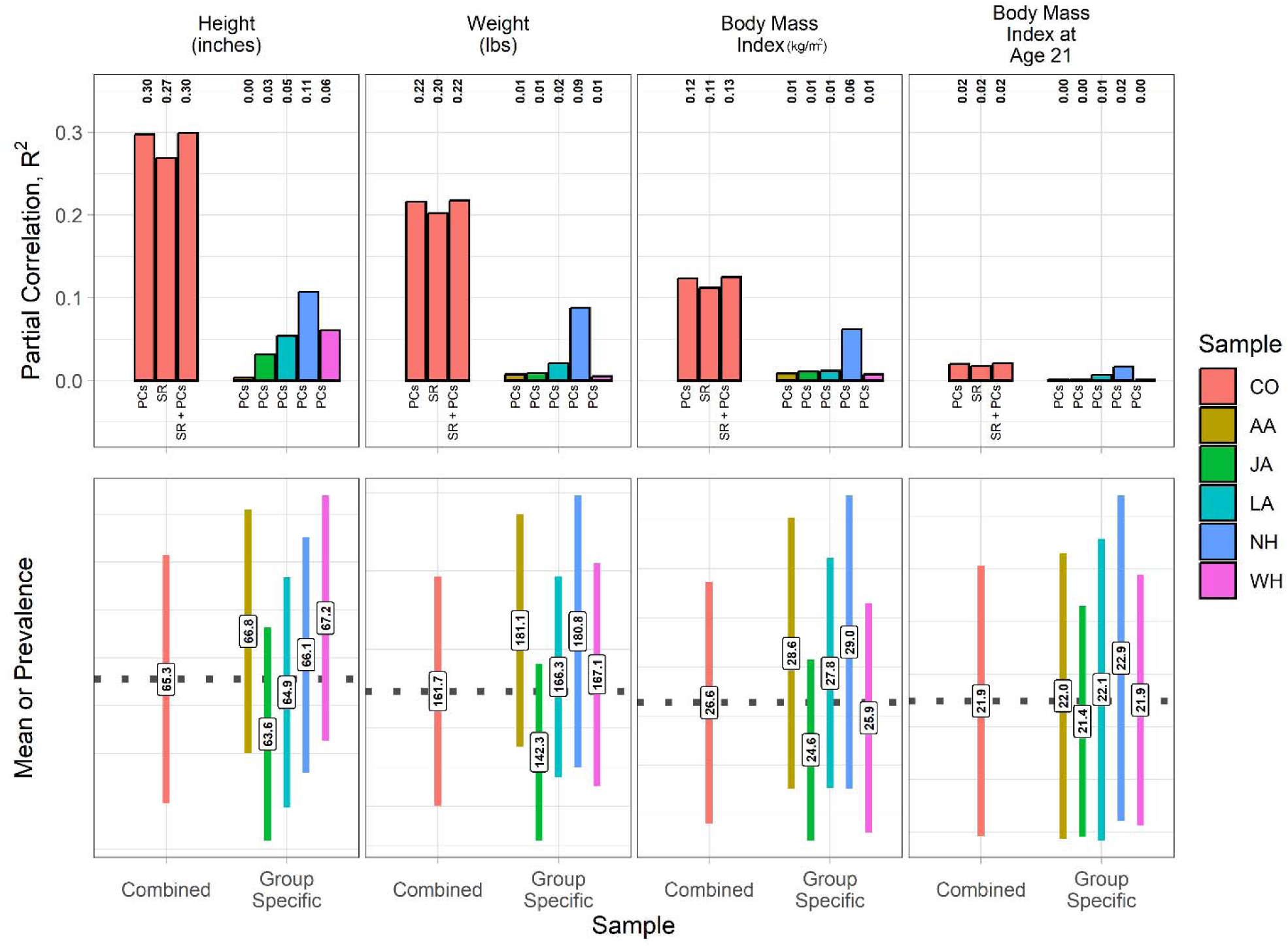
Partial Correlations of Anthropometric Measures. CO = Combined, AA = African American, JA = Japanese American, LA = Latino, NH = Native Hawaiian, WH = White

**Supplemental Figure 16:**
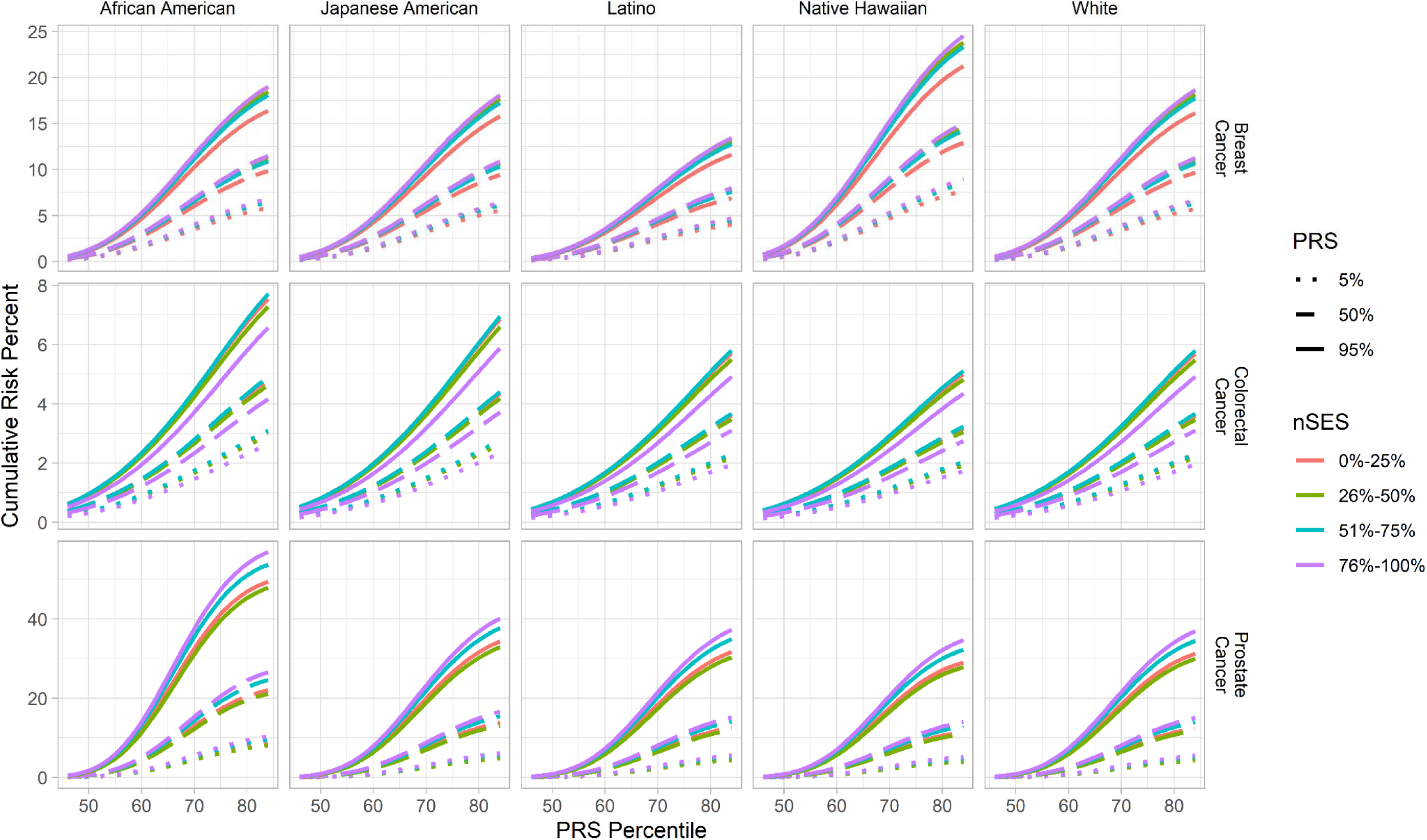
Cumulative risk of cancer by PRS and nSES

**Supplemental Figure 17:**
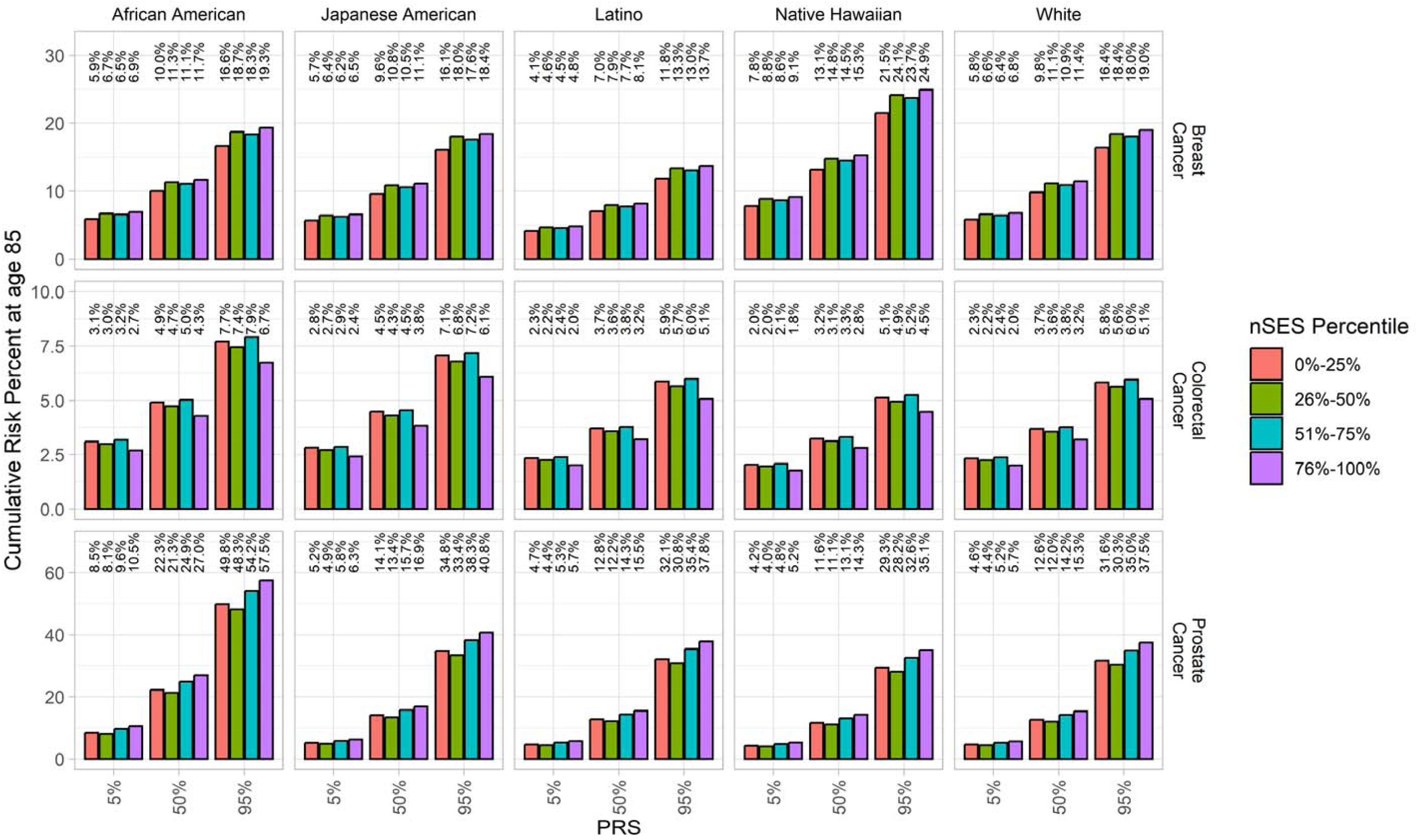
Cumulative risk of cancer by PRS and nSES at age 85

**Supplemental Figure 18:**
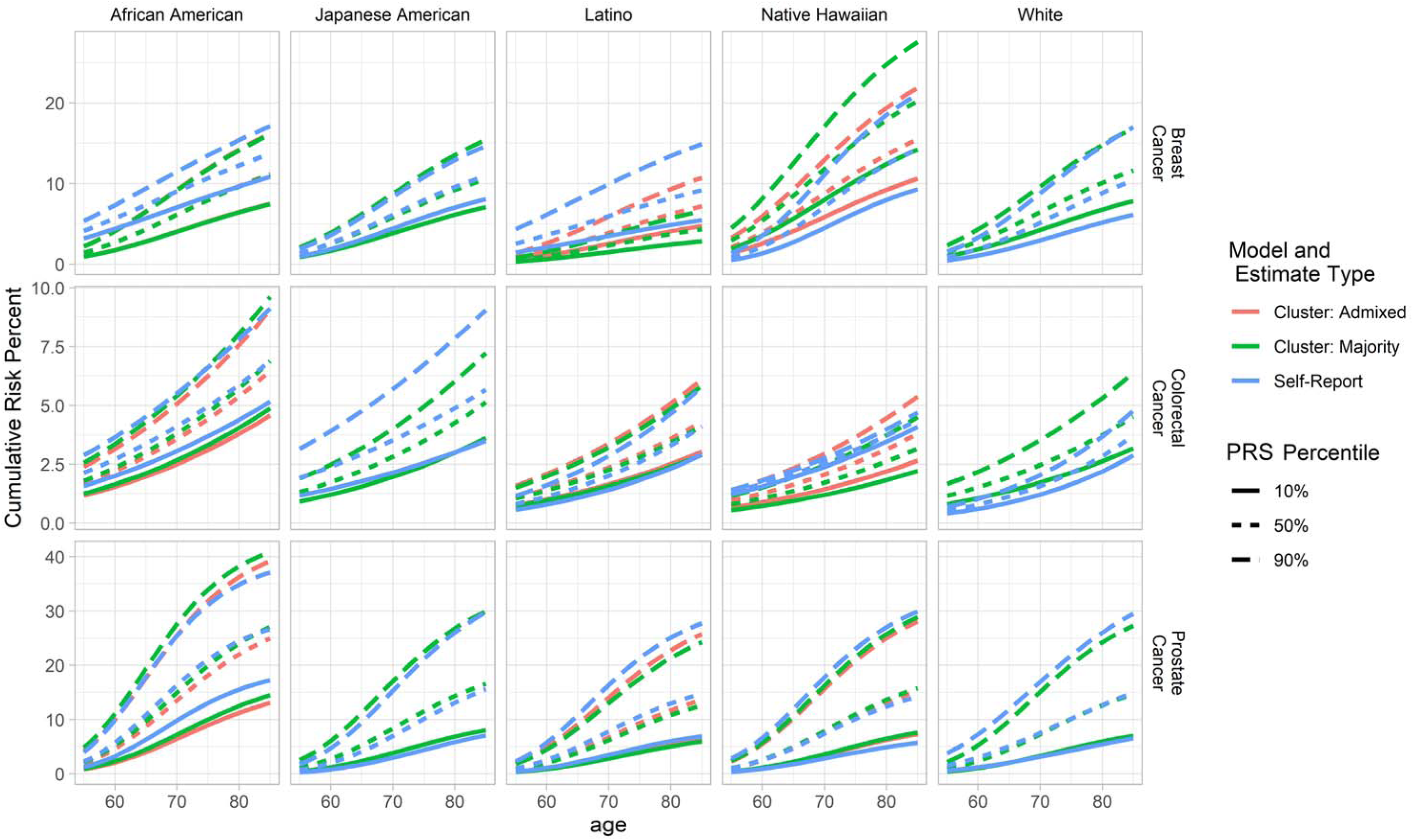
Cumulative risk by cluster values and self-reported race and ethnicity

**Supplemental Figure 19:**
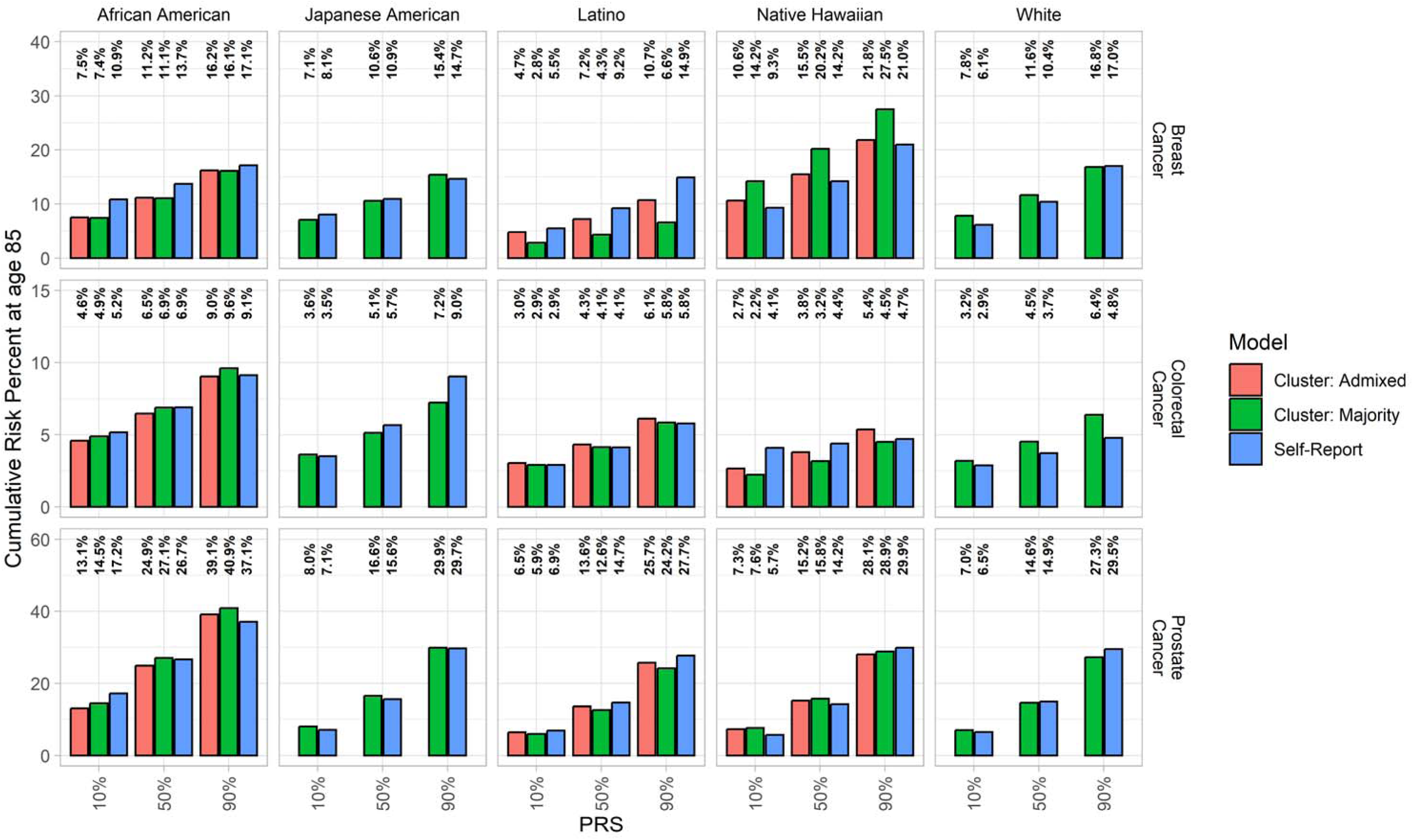
Cumulative risk by cluster values and self-reported race and ethnicity at age 85

### Supplemental Text

#### Race and Ethnicity Group Assignment Algorithm

Cohort participants had the option to select multiple racial groups in additional to self-reported ethnicity (defined as Hispanic or non-Hispanic origin). Most MEC analyses categorize participants into one of the five major racial and ethnic groups that were targeted during recruitment; however these detailed data may be used in description of these cohorts. In the case of participants self-reporting multiple racial groups, in addition to Hispanic origin, participants were preferentially assigned to racial and ethnic groups based on the following order: African American (also coded as Black), Native Hawaiian, Latino (also coded as Hispanic), Japanese American, White. For example, if an individual reports white and Native Hawaiian, they would be classified as Native Hawaiian for these analyses.

For illustration of this data source, we used the grouping of self-reported race and ethnicity outlined above to concisely describe general database characteristics. Although these groupings provide a simplified summarization of the data, there are inherent limitations of terminology that reduces genetic, cultural, and lifestyle variation into self-defined groups. The MEC and all data sub-sets contain more detailed information on multiple self-declared racial and ethnic group memberships, paternal and maternal group memberships, “write-in” responses, participants’ country birthplace, maternal/paternal birthplaces, and a wide range of dietary, lifestyle, environmental, and genetic variables which may better explain variation in disease risk than self-identified group membership. For detailed self-reported racial and ethnic group used in supplemental principal component figures, all races responded to are included in the order: Black, Hawaiian, Latino, Japanese, White, Filipino, Chinese, Korean and Other. If the person is Chinese-Hawaiian-Samoan they would have the value of “HCO” for this variable. This information is available for all current and future analyses through data requests.

#### Quality Control

The Multiethnic Cohort (MEC) genetic repository includes over 30 genome-wide association study projects conducted since 2009 using various Illumina genotyping arrays [Supplemental Table ST1]. Certain projects that utilized the same or similar Illumina arrays, primarily for MEGA chip and OncoArray projects, were merged into a single dataset. Imputations were performed by project and racial/ethnic group through the TOPMed imputation server (https://imputation.biodatacatalyst.nhlbi.nih.gov/).

##### GWAS Quality control (QC) procedure

Each GWAS project has been cleaned either as part of a larger dataset or by itself. QC steps for variants include call rate >= 0.95 and one or more of the following filtering criteria:

1. minor allele frequency
2. concordance rate of QC replicates
3. HWE p-value
4. Illumina GenomeStudio Gentrain score or separation score
5. poor clustering by visual inspection
6. mendelian errors in HapMap trios or duos
7. sex difference in allele frequencies for autosomes/XY
8. sex difference in heterozygosity >0.3 for autosomes/XY
9. positional duplicates
10. not in the 1000 genomes reference panel
11. frequency outliers when comparing to ethnic-specific 1000 genomes reference subjects
12. allele mismatches when comparing to the 1000 genomes reference panel

QC steps for subjects include call rate >= 0.95. Cryptic replicates based on genotype data have been investigated and removed if there is an indication of sample error. A very small number of cryptic replicates (∼23) are retained because their questionnaire data suggests they may be potential twins. Genotypes for known replicates across projects have been compared to each other to ensure they correspond to the same individual. Gender checks have been performed by evaluating either the mean intensities of chromosome X and Y probes, heterozygosity for X chromosome variants, or both. Subjects with sex mismatches in relation to their questionnaire data are excluded.

Additional QCs have been conducted for merging multiple MEGA and OncoArray projects respectively. In total, there are five mega chip projects using three slightly different MEGA arrays from Illumina (MEGA_Consortium_v2_15070954_A2, MEGA_Consortium_v1-1_15071648_A1 and MEGA_Consortium_15063755_B2), along with seven OncoArray projects.

1. Make sure to start with a dataset without MAF filtering for each project
2. Ensure that alleles in each dataset are on the genome forward strands (or at least consistent across projects), and that subjects have been filtered by a call rate >= 0.95
3. Filter out variants with a call rate < 0.98 in each dataset
4. Limit the variants to the overlapping variants across all the datasets
5. Exclude variants with replicate concordance < 1 based on cross-project subject pairs (740 on Mega Array and 179 on Oncoarray)
6. Exclude replicate subjects with a lower call rate
7. For African American, Latino, Japanese American and White populations, compare their allele frequencies to ethnic specific frequencies of 1000 genomes reference subjects, and filter frequency outliers (frequency difference >= 0.2 or 0.25)

#### Imputation

The cleaned GWAS datasets were initially in the GRCh37 genome build. The triple-LiftOver was used to convert their positions to GRCh38. Additionally, for SNPs situated in Between-Builds Inverted Sequence (BBIS) regions identified during triple-LiftOver, their strands were flipped to ensure consistency with the reference panel in the new genome build. Subsequently, these datasets were submitted to the TOPMed imputation server (https://imputation.biodatacatalyst.nhlbi.nih.gov/) for QC and imputation. The Imputation pipeline used ranges from michigan-imputationserver-1.6.0 to 1.6.6, the imputation software was minimac4-1.0.2 and the phasing software was eagle-2.4.

##### High-Dimensional Data Tabulation

To fit Poisson time-to-event models in genic risk score analyses, we produced finely tabulated lifetable data from individual-level data from the MEC Genetics Cohort. To generate this data, we stratified individual-level data by categorical variables (race and ethnicity [5 levels] and categorical polygenic risk score defined by evenly distributed cut-points [6 levels]. Within stratified data we tabulated number of cases and person-years across 5-year categories, then within each stratum we calculated mean polygenic risk score and age, weighted by an individual person-years contribution to each stratum.

In this time-to-event analysis, person-years begins to accumulate at age of biospecimen collection, not initial MEC cohort enrollment. Although germline variation does not change over the study period, contributing a biospecimen and participation in the genetics cohort, requires participants to survive many years to sample collection. This interval of time between cohort enrollment and sample collection would represent immortal person-time, where it would be impossible for participants to experience death, and inclusion of this time would produce incorrect incidence estimates.

##### Absolute Risk Calculation

We estimated absolute risk, in the form of cumulative and 5-year risk, using Poisson regression models fit for each cancer type as an outcome across each cancer-specific dataset. To account for all-cause mortality as a competing event, we fit the same models with all-cause mortality as the outcome. Using these two models, we estimated cumulative risk as follows:

Age-specific cumulative risk within risk strata *k* is 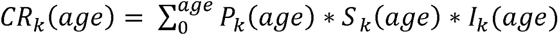.

- *K* represents a covariate level defined by vector of variable levels. For example, *k = (PRS) =* (0) represents an individual with a standardized PRS value of 0 (i.e. 50% PRS risk). To estimate at alternative risk percentiles, we used the standardized PRS distribution to identify standardized PRS values that correspond to each quantile, then used the standardized PRS value in a linear combination to estimate expected incidence and/or risk.
- *P_k_(age)* is the probability of survival from all-cause mortality up to a specific age, within strata *k*, estimated using the all-cause mortality hazard model. The hazard is transformed to survival probability using the formula 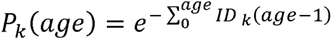, where *ID* is age and exposure level (*k*) specific hazard of all-cause mortality.
- *S_k_(age)* is the probability of not experiencing the specific cancer at an earlier age up to a specific age, within strata *k*, estimated using the cancer hazard model. The hazard is transformed as in the mortality model but now using the cancer model, 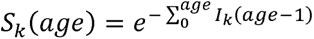, where *I_k_* is age and exposure level (*k*) specific hazard of cancer.
- *I_k_(age)* is age and exposure level (*k*) specific hazard of cancer.

Estimation of 5-year risk is done using a modified version of the above formula, where cumulative risk in an interval is (age, age+5) is conditional on survival up to the given age before cumulative risk within the interval is accumulated.

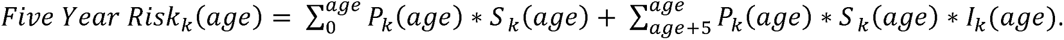

##### Absolute Risk Error Estimation

Error of each absolute risk estimation was calculated using Monte Carlo resampling of beta coefficients. This method was implemented by taking fit Poisson regression estimates for both cancer and all-cause-mortality models, then sampling new beta terms from a normal distribution using the initial beta as a mean and the initial standard error as the variance. Following the resampling we estimate both cumulative and 5-year risk as above. We completed this resampling procedure 5000 times to create an absolute risk and 5-year risk estimate distribution across all ages and exposure levels. The 95% confidence interval bounds represent the 2.5% and 97.5% values of the distributions.

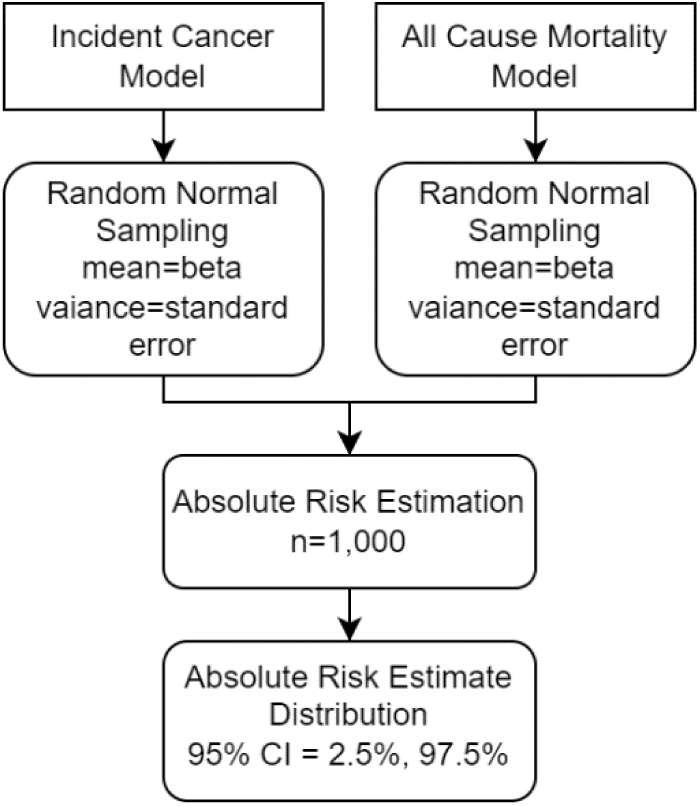

## Supplemental Tables

**Supplemental Table 1:**
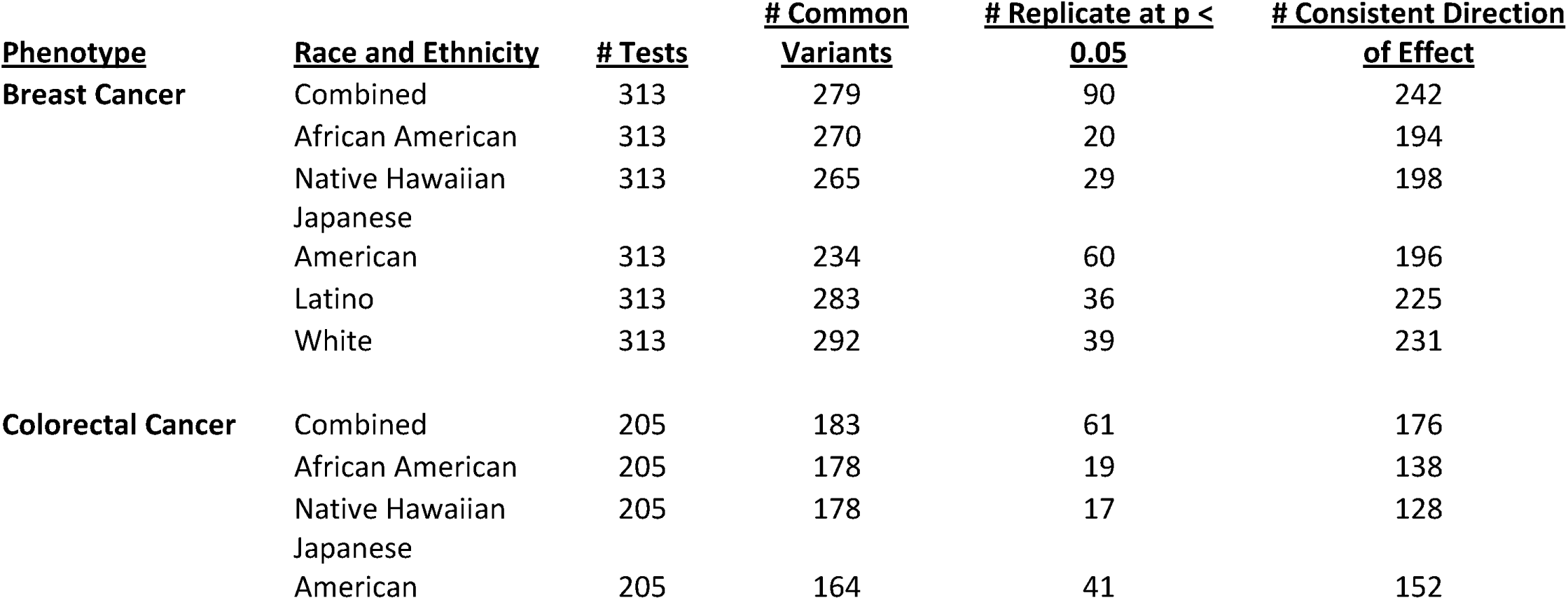
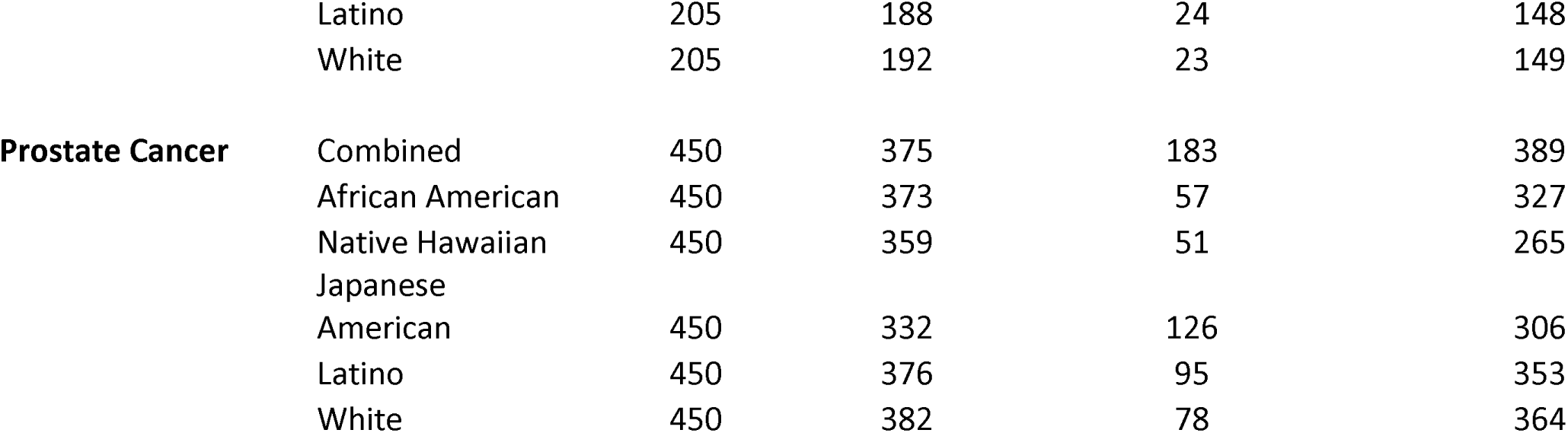
Replication Summary Statistics.

**Supplemental Table 5:**
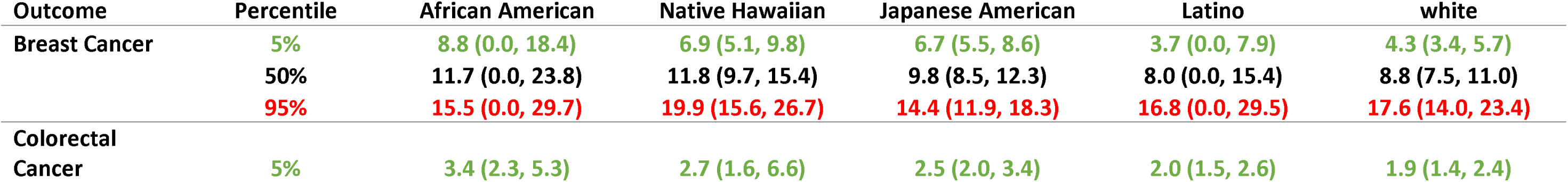
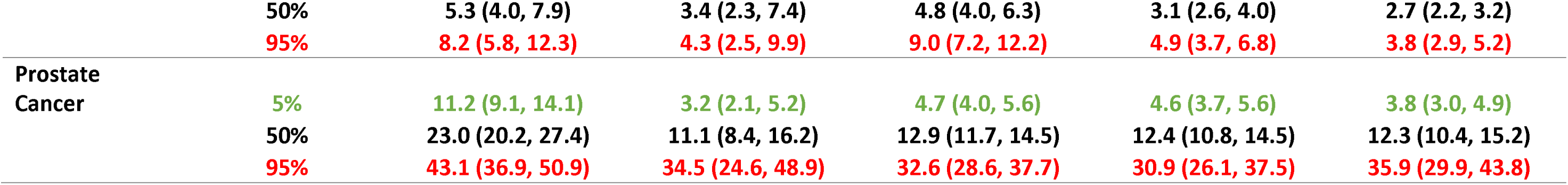
Cancer and PRS Specific Cumulative Risk (as percentage) at Age 80.

